# Patient Flow in Congested Intensive Care Unit /Step-down Unit system: Premature Step-down or not?

**DOI:** 10.1101/2022.12.16.22283534

**Authors:** Yawo M. Kobara, Felipe F. Rodrigues, Camila P. E. de Souza, David A. Stanford

## Abstract

A Step-Down Unit (SDU) provides an intermediate Level of Care for patients from an Intensive Care Unit (ICU) as their condition becomes less acute. SDU congestion, as well as upstream patient arrivals, forces ICU administrators to incur costs, either in the form of overstays or premature step-downs. Basing on a proxy for patient acuity level called the ‘Nine Equivalents of Nursing Manpower Score (NEMS)’, patients were classified into two groups: high-acuity and low-acuity. Two patient flow policies were developed that select actions to optimize the system’s net health service benefit: one allowing for premature step-down actions, and the other allowing for patient rejection actions when the system is congested. The results show that the policy with patient rejection has a net health service benefit that significantly exceeds that of the policy with the premature step-down option. Based on these results, it can be concluded that premature step-down contributes to congestion downstream. Counter-intuitively, premature step-down should therefore be discouraged and patient rejection actions should be further explored as viable options for congested ICUs.

## 1. Introduction

Intensive care units (ICUs) provide care to patients with high levels of acuity. Patient acuity has often been measured by standardized scores such as SOFA and the variants of APACHE (Knaus et al. 1981; Lambden et al. 2019) as well as nursing manpower scores, such as the “Nine Equivalents of Nursing Manpower” (NEMS) score (Perren et al. 2012; Miranda et al. 1997). Staffing is typically one nurse per ICU patient, and ICU beds are rarely idle. During recovery, the continued needs for intensive care (and consequently, an ICU bed) diminish, and this is reflected in a lower NEMS score. To provide better continuity of care, so-called Step-down Units (SDUs) are intended for these recovering patients (Lekwijit et al. 2020; Lu et al. 2014; Gershengorn et al. 2020), with staffing of typically one nurse per two SDU patients. In a congested setting, patients with lesser acuity who continue to occupy ICU beds represent at best a sub-optimal use of resources, and at worst may prevent an arriving high-acuity patient from getting the care she requires.

The NEMS score is a scoring derived by Miranda et al. (1997) from the therapeutic intervention scoring system (TISS) to determine the required levels of intensive care needs, provide information on the severity and prognosis of patients’ acuity and determine the number of nurses needed and their workload. The NEMS score is a value between 0 and 56 points and represents the sum of nine (9) patient related factors (see Table: 2) that influence nurses’ workload as they administer care (Carmona-Monge et al. 2013; Miranda 1997; Vuković 2020).

ICU beds and staffing represent a high operating cost for any hospital (Halpern and Pastores 2015; Wunsch et al. 2012; Seidel et al. 2006). Therefore, with its highly valued care and increasing demand, the ICU must improve its flow to optimize system throughput (Armony et al. 2018). For a patient to be discharged or transferred from the ICU, a physician’s declaration of the patient’s medical stability is required (Nates et al. 2016; of Critical Care Medicine of the Society of Critical Care Medicine et al. 1999). Determining a patients’ suitability to leave the ICU often takes time. Moreover, if no beds are available downstream, patients may occupy ICU beds longer than medically necessary (Stelfox et al. 2018). Furthermore, ICUs avoid rejecting patients because there is often a risk of death if a patient is turned down or left untreated. As such, patient arrivals may trigger the step-down of a suspected lower acuity patient to free up a bed for the recent arrival. Motivated by these phenomena, this study defined two types of ICU patient transfers or ‘step-downs’: regular step-down, during which sufficiently low-acuity patients are moved from the ICU to the SDU and premature step-down, during which a high-acuity patient is moved from the ICU to the SDU before reaching her intended medical stability.

In the case of increasing demand for intensive care and a congested ICU, management may need to decide between rejecting a new patient in need of critical care and prematurely stepping down a current occupant. Possible future scenarios, including demand surges due to new diseases, may put the ICU in a precarious situation. As an illustration, a rise in the number of coronavirus disease 2019 (COVID-19) cases may result in a spike in the demand for hospital admission and critical care. In such a situation, when the ICU is full, and resources are constrained who receives the service: the newly arriving patient or the existing patient (Azcarate et al. 2020)?

This study compared two policies governing patient flow. The first combined the following actions: reject or admit an arriving high-acuity patient to the ICU, step-down or an retaining existing low-acuity patient in the ICU, premature step-down or retain a high-acuity patient in the ICU, and prematurely discharge or retain a low-acuity patient from the SDU. In the second policy, however, whenever the system is congested and a patient arrives, an existing high-acuity patient is prematurely stepped down to admit the arriving patient instead of rejecting her as would happen in the first policy. In this policy, premature step-down of an existing patient and admission of an incoming patient has priority over rejection of the latter under congestion. The variation in the system’s health service benefit in a congested environment is quantified using a metric that reflects the benefit or detriment of an action, i.e., an action either increases or decreases the system’s health service benefit. The aim is to assess the impact of the two decision policies on patient flow in a congested environment. The main difference between the two policies is that the first performs premature step-downs of existing high-acuity patients to avoid rejection of arriving patients when the ICU is full, whereas once the ICU is full, the second rejects arriving patients. We sought to optimize the long-term health service benefit of these policies. In the methodology used, relative weights were assigned to each atomic action, a Markov decision model was built and solved to obtain optimal actions that made up a policy based on such weights. The optimal actions of the two policies were then analyzed for sensitivity and used as inputs to simulate the hospital management flow and compare the two policies under an increasing rate of arrival.

The remainder of the paper proceeds as follows. Section 2 provides an overview of related literature, with keen attention to the application of operations research to patient flows in the ICU. Section 3 briefly describes the empirical data used in this research. Section 4 describes the simple solution methodology of the infinite horizon Markov decision process model proposed in this paper, its sensitivity analysis, and the simulation model. Section 5 presents the result of the decision rules for each policy, a sensitivity analysis, and the simulation results. Section 6 discusses the implication of the results obtained, and the paper closes in Section 7 with conclusions and recommendations.

## 2. Overview of Related Literature

ICU Patient flow and capacity planning have received lot of attention in the operational research literature, even more so during the COVID-19 pandemic (Shoukat et al. 2020; Chin et al. 2020; Moghadas et al. 2020). Several papers have addressed patient flow and capacity planning based on resources such as beds and staffing and their scarcity. Comprehensive reviews of this literature can be found in Bai et al. (2018); Lin et al. (2009). The present review focuses on a few key papers in the literature that address decision-making in an ICU under congestion. The ICU flow decision-making process is complex and challenging. Azcarate et al. (2020) remark that flow decision guidelines in the ICU are hindered by the lack of clear objective metrics to determine the patients that are likely to benefit from remaining in critical care. Levin and Charles (2001) observed that only a small number of ICUs have developed formal patient discharge guidelines. Most ICUs follow empirical decision processes and rely on consensus, as opposed to scientific evidence, which highlights the importance of patient flow policies.

In a congested scenario, it may be customary to triage current ICU patients using the (Sprung et al. 2013) method. Much like Levin and Charles (2001), most of the literature recommends patient discharge as a means of reducing ICU overcrowding. Delays or overstays in discharge may also result from bed shortage in a hospital’s downstream units. Lack of beds in the cascade or disagreements about admitting services in the wards were the main causes of failed ICU discharges (Silva et al. 2014; Levin and Charles 2001; Armony et al. 2018).

Downstream congestion has also been observed to cause blockage in the ICU, which keeps patients from moving (Cochran and Bharti 2006). Mathews and Long (2015) found that in the United States, ICU patients who are ready for transfer to a downstream unit often stay in the ICU for longer than clinically necessary. Most such patients remain in a critical care bed and thereby delay admission for other incoming patients. In their studies, Mathews and Long (2015) examined different discharge policies under of capacity constraints in the Emergency Department (ED). Shi et al. (2016) developed a stochastic network queuing model with dynamic discharge policies for peak utilization. Their model proved to reduce admission delays as well as ED wait times for admission to the ward.

Markov Decision Process (MDP) models have been used actively in recent years in hospital resource and inventory management in general, and ICU resources and service modelling in particular. (Broyles et al. 2011; Dobson et al. 2010; Patrick et al. 2008; Patrick 2012; Chan et al. 2012; Li et al. 2018; Nunes et al. 2009) all used discrete-time MDP to model discharge and admission decisions in the ICU with many dissimilarities in the resulting models. One major difference is the state space definition of the MDP model.

For effective hospital resource usage, Nunes et al. (2009) suggested an MDP model for elective (non-emergency) patient admissions. The model’s goal is to avoid wastage and excessive usage of resources. It was discovered that an ideal admission control approach kept resource use near to targeted utilisation levels. However, the enormous complexity and complicated stochastic dynamics of the model made application challenging.

Patrick (2012) also developed an MDP model to examine various patient and physician lead time scenarios. They used simulation to show that the MDP model performs effectively across a wide range of conceivable circumstances.

Chan et al. (2012) studied priority demand-driven ICU discharge strategies to determine their effects on patient mortality and the overall readmission burden. To reflect the overall occupancy of the ICU, they defined the status as the numbers of various kinds of patients. They created an approximation method and discovered the best practices for particular regimes. Their admissions choices were examined using an MDP methodology by Li et al. (2015). Their notion of a state was comparable to that of Chan et al. (2012). However, they coupled the number of various patient in the ICU with the number of beds that were available. To analyze and enhance the admission policy upper and lower bounds of the parameter were determined.

To quantify the effect of the number of reserved beds and recommend when to prematurely release existing patients, Li et al. (2018) established the analytical framework of an MDP model with the system state described by the numbers of two kinds of patient in the ICU. The model was designed to strike a balance between rejection of entering patients and premature discharge, after comprehensive numerical experiments to examine the influence of each parameter on total survival benefits.

Metrics have been developed by the clinical community as systematic criteria to evaluate patient health severity status. Rodrigues et al. (2018), working with a large dataset from an academic hospital, used a discrete event simulation to show the benefits of SDU beds in optimizing hospital expenses and patient flow at a jam-packed facility and developed a metric called the Nine Equivalents of Nursing Manpower (NEMS). Shmueli et al. (2003) used Acute Physiology and Chronic Health Evaluation II (APACHE II) to evaluate patient severity. The NEMS and APACHE are based on clinical observation of patients and are generally assigned daily based on data available during an ICU stay. Strand and Flaatten (2008) provided a review of several versions of three prognostic scoring systems to review severity metrics in the hospital. Kim et al. (2015) estimated the cost of denied ICU care for all the medical patients admitted to 21 hospitals through the EDs. They empirically found that ICU congestion could have a significant impact on ICU admission decisions and patient outcomes.

The step-down and discharge process in the ICU-SDU system is currently prolonged beyond the acute care days of patients. The unavailability of empty beds in the SDU results in patients using the ICU even when they have no need of it (Armony et al. 2018; Lekwijit et al. 2020). Congestion in the SDU contributes to that in the ICU and therefore produces an increased length of stay. When patients who do not need the ICU service stay in the ICU longer than needed, this prevents the admission of others who need it the most and exposes them to higher risk of mortality. The question worth asking here is what the hospital can do in terms of step-down policy planning, not only to reduce length of stay, but also to increase the health service benefit of patients who request ICU care. To answer this question, the first step is to consider how the step-down process is currently performed.

Studies on the impact of SDUs are primarily limited to observational and simulation-based models with different objectives (Mcilroy et al. 2006; Doolan et al. 2005; Richards et al. 2012; Prin and Wunsch 2014). Most studies to date recommend SDUs as a safe care option for patient who does not need ventilation (Prin and Wunsch 2014). However, these studies have not been conclusive on the benefit of the SDU in reducing mortality. Many hospitals have used SDUs specifically as an alternative to full intensive care and this practice is thought to be an alternative level of care. Continued research and data collection from the SDU is needed and required in this arena to contribute to the development of the patient discharge process to characterize and completely specify the medical and physiological step-down as SDU policy, and to enable for comparison of outcomes across different units.

In the literature, few research papers have used MDP to model ICU patients flow (Bai et al. 2018). Those that did, determined only one aspect of the flow. Chan et al. (2012) determined an occupancy threshold to perform premature discharge. Li et al. (2015) and (Chan et al. 2012) focused on an ICU occupancy threshold to initiate rejection and/or premature step-down, but their model considered only the ICU. Li et al. (2018) developed a model to quantify the feasible number of beds to reserve in the ICU to start rejection and premature discharge of lower-acuity patients. The objective here is to determine the optimal decision under congestion.

This paper looks at congestion from the last-bed problem perspective. When the ICU is full how to decide between rejecting a new patient in need of critical care and creating a vacancy by prematurely discharging a current occupant? Azcarate et al. (2020) offer a review of literature on this clinical management dilemma with factors to consider and the patient health consequences of each decision. However, they noted that mathematical models of ICU management practices overlook these health factors and their consequences to patients. In response to the existing literature, this study proposes to determine actual decisions to be made in congestion instead using a certain threshold considering risk factors. Decisions about all aspects of patient flow are considered instead of focusing on a subset. The authors suggest focusing on the congested area instead of solving a bulky state space with states that are not relevant to congestion. Finally, the simulated results obtained here counter-intuitively suggests that rejection of high-acuity patients when the system is full is better than to the conventional practice of prematurely stepping down another high-acuity patient.

## 3. Data Description

Empirical data from the London Health Sciences Centre (LHSC) have been used to estimate the distribution function of the transition used for the simulation. The LHSC is a multi-site health care facility with two main hospitals: University Hospital and Victoria Hospital, which includes the Children’s Hospital at London Health Sciences Centre. The data used were a set of four-year records containing more than 70000 logs with nearly 8000 patients from January 2015 to December 2018. The Patient information includes patient age, gender, admitting diagnosis, admitting source, discharge destination, and daily NEMS scores until discharge. The NEMS and patient health are closely associated because as the patient’s health improves, less nursing care is required, which lowers the NEMS. Empirically, a patient is deemed to have “Very low-acuity” if their score is less than 10, “low-acuity” if their score is between 11 and 25, and “High-acuity” if their score is between 26 and 56 (See Table 1) (Rodrigues et al. 2018). From Figure 1, it is observed that more than 95 % of patients requesting the ICU’s services have an NEMS score higher than 25. It is therefore reasonable to assume that all patients requesting the ICU are high-acuity patients.

**Table 1.** Severity and Levels of Care Characteristics. Source: Rodrigues et al (2018)

| Level of Care | Beds characteristics | Patient per nurse ratio | Cost (\$ /patient-day) | NEMS Score |
| --- | --- | --- | --- | --- |
| 1 | Standard Ward beds:<br>No organ support, no ventilation | 3 or more to 1 | \$600 | $\leq 10$ |
| 2 | Step-down beds: Support single failed organ system, no ventilation | 2 to 1 | \$2,000 | 11 to 25 |
| 3 | Intensive care beds:<br>multiple organ support | 1 to 1 | \$3,500 | 26 to 56 |

**Figure 1.**
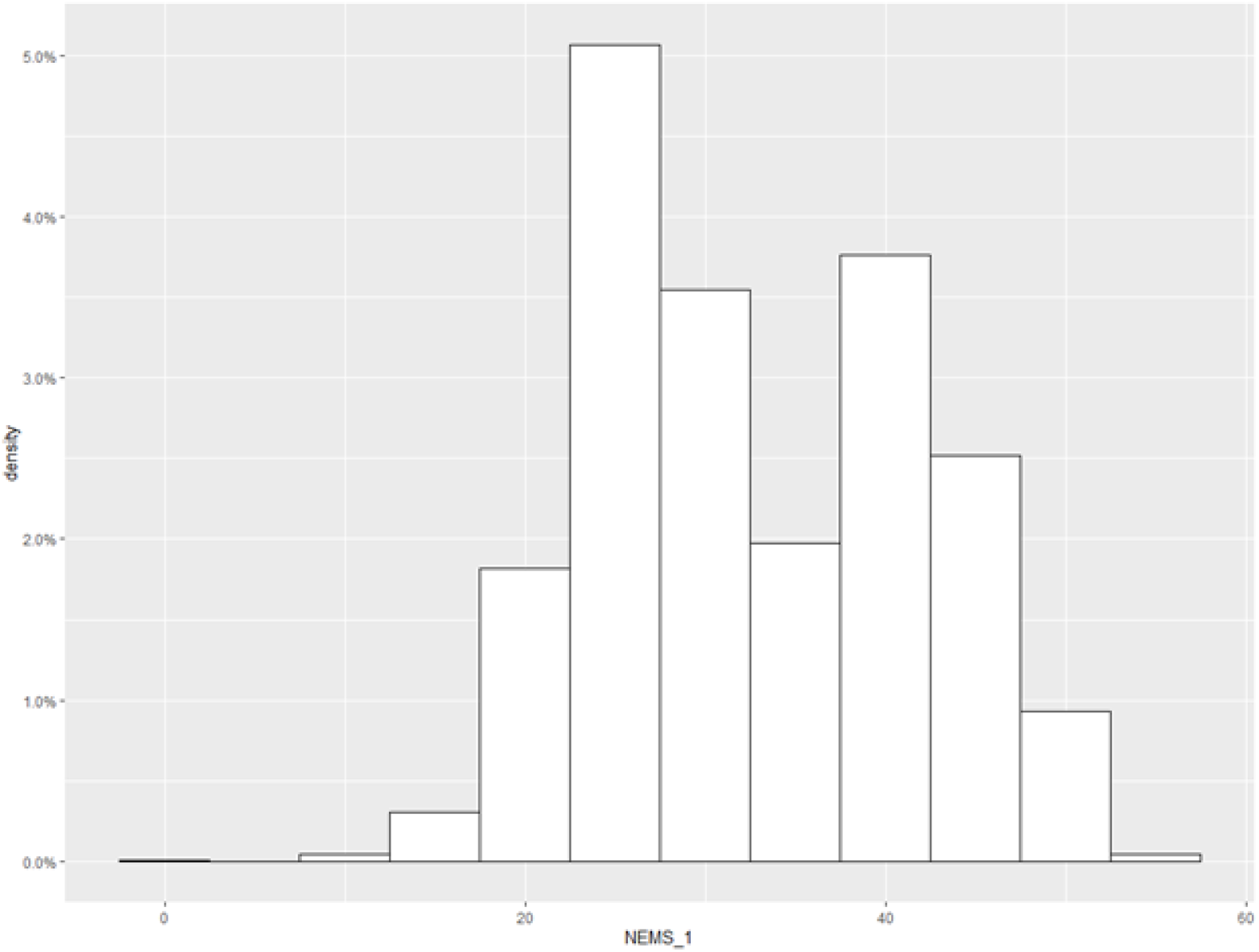
First-day NEMS scores of patients in system

Patients arrive at the ICU individually from different sources. The ED, a unit with varying patient severity provides the highest proportion of ICU patients. From the hospital studied, about 38% of the patients come from the ED, 22% from the ward, 21% from the Operating room, and 20% from other places such as other hospitals or the SDU. 99.1% of admissions into the ICU are unplanned and are patients requiring immediate medical care. With the priority triage policy used in many hospitals, the hospital studied has very little, or no control over admitting high-acuity patients arriving through the emergency route. Daily arrivals are essentially equally distributed with Thursdays having the maximum number of admission. The hourly admission trend was also examined. Figure 4 show the inter-arrival time distribution of the patients. The average inter-arrival time is around 6.47 hours and is approximately exponential. The system’s capacity is 30. Figure 2 is the daily ICU occupancy in 2018. There is no evident trend in the data. Nevertheless, occupancy is observed to be often higher than capacity. Congestion is a daily routine. In most cases, to offset overcapacity, patients are placed elsewhere at an alternative level of care (ALC). Figure 3 shows a time plot of the various acuity levels observed in the system. The evolution of patients’ acuity levels depends on the severity of their condition and the care they receive.

**Figure 2.**
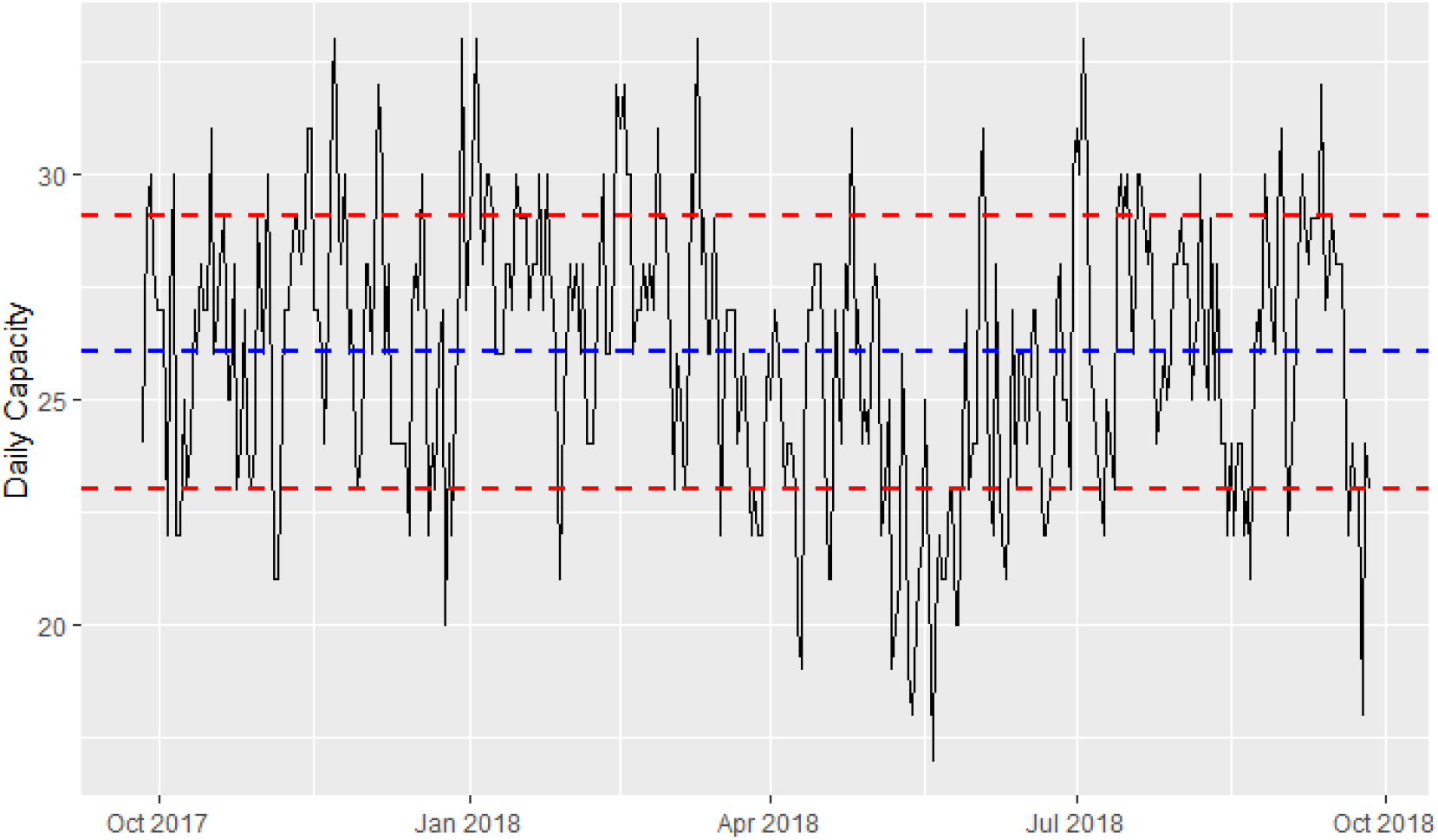
Time plot of daily occupancy in 2018. (Blue line represents the mean and the red lines one standard deviations from the mean.)

**Figure 3.**
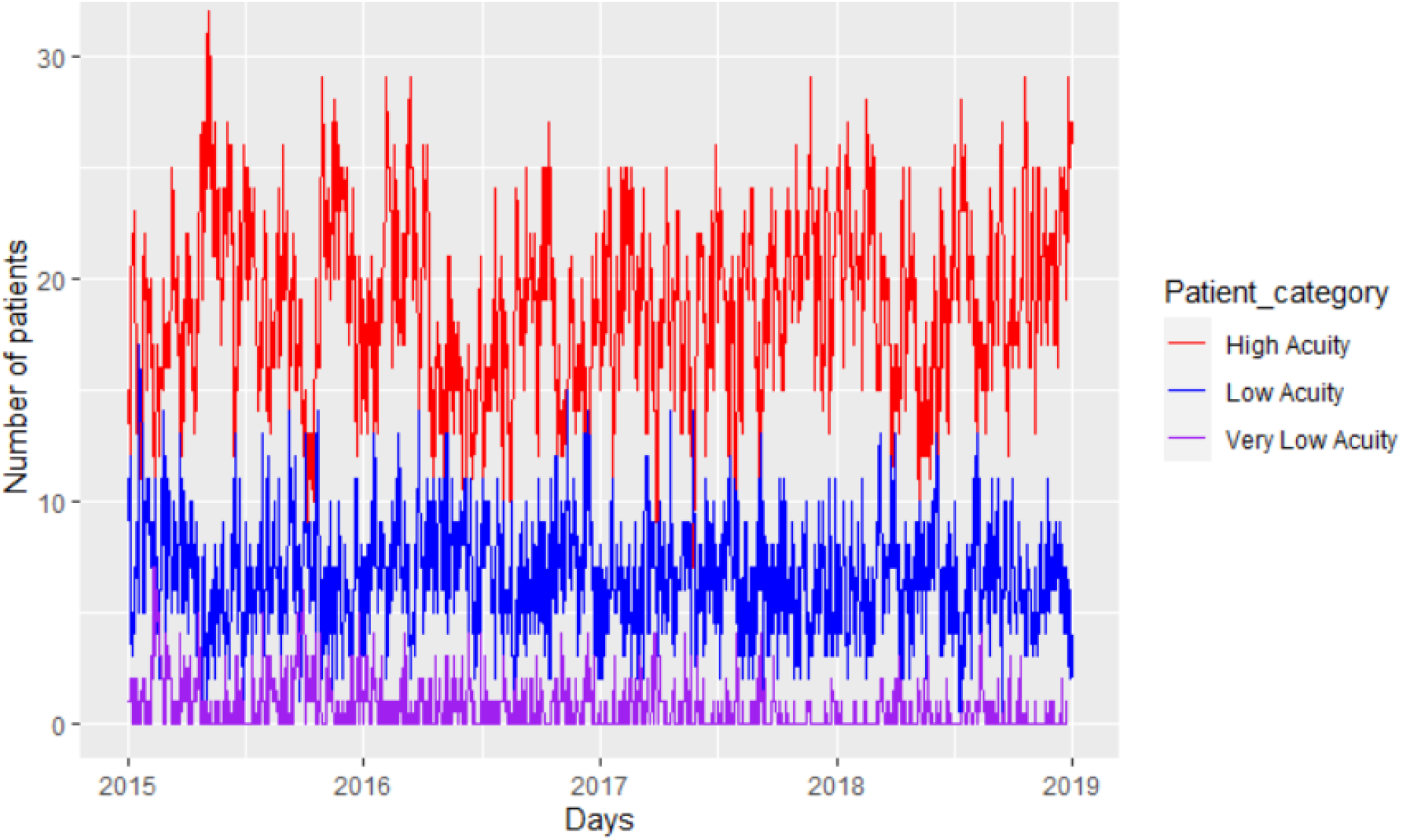
Time plot of daily occupancy by acuity levels’.

**Figure 4.**
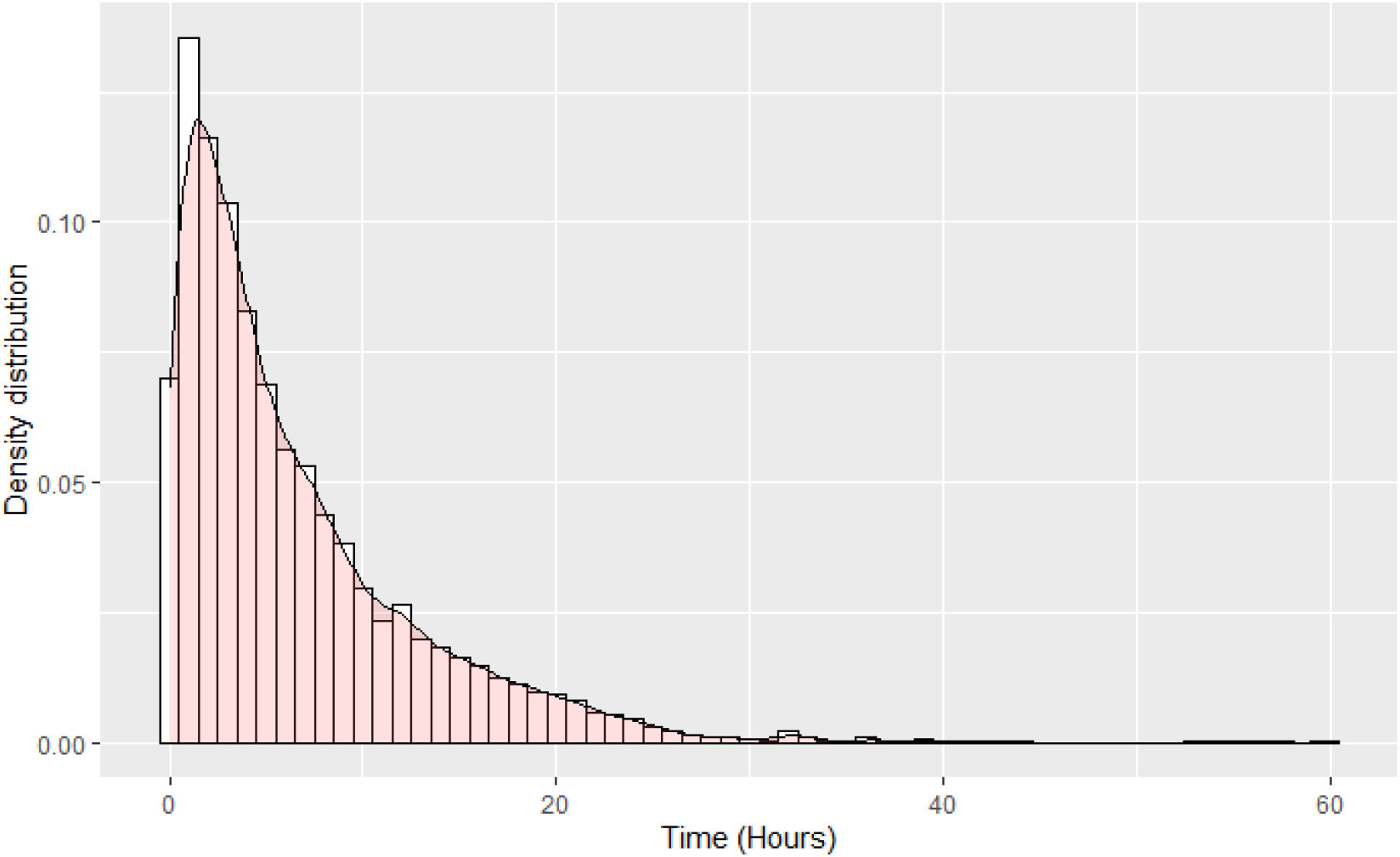
Density distribution of system inter-arrival time (Blue line represents the mean and the red lines one standard deviations from the mean.)

The daily number of the various categories of patients in the ICU was recorded. From that, the distribution of the number of patients that recovered from one acuity level to the other in the system was obtained. Figure 5 presents the density function of the number of patients that recovered from high-acuity to low-acuity at the ICU. These were patients with NEMS scores between 11 and 25 that were destined to move from the ICU to the SDU. Figure 6 displays the density function of the number of patients that recovered from low-acuity to largely recovered in the system. These were patients with NEMS Scores less than 10 who were moved from the SDU to the general ward. The patients who died or left the ICU directly to home were called discharged. Because the number of recovered patients is countable, the number of occurrences is independent, and the estimated average rates of every occurrence are approximately equal and independent it can be safely assumed that the recovery processes follow a Poisson distribution with parameter estimates 2.45 and 3.22 for high-acuity and low-acuity recoveries respectively. Table 3 provides the descriptive statistics of the recovery processes.

**Table 2.** NEMS Components (*Source*:*Mirandaet al*. (1997))

| Items | Points |
| --- | --- |
| 1. Basic monitoring: hourly vital signs, regular record and calculation of fluid balance | 9 |
| 2. Intravenous medication: bolus or continuously, not including vasoactive drugs | 6 |
| 3. Mechanical ventilatory support: any form of mechanical/ assisted ventilation, with or without PEEP | 12 |
| 4. Supplementary ventilatory care: breathing spontaneously through an endotracheal tube; supplementary oxygen by any method, except if (3) applies | 3 |
| 5. Single vasoactive medication: any vasoactive drug | 7 |
| 6. Multiple vasoactive medications: more than one vasoactive drug, regardless of type and dose | 12 |
| 7. Dialysis techniques: all | 6 |
| 8. Specific interventions in the ICU: such as an endotracheal intubation, the introduction of a pacemaker, cardioversion, endoscopy, emergency operation in the past 24 h, gastric lavage; routine interventions such as X-rays, echocardiography, electrocardiography, dressings, the introduction of venous or arterial lines, are not included | 5 |
| 9. Specific interventions outside the ICU: such as surgical intervention or diagnostic procedure; the intervention/procedure is related to the severity of illness of the patient and makes an extra demand upon manpower efforts in the ICU | 6 |

**Table 3.** Descriptive statistics of the recovery process

|  | Mean | Var | Median | Min | Max | Skew | Kurtosis |
| --- | --- | --- | --- | --- | --- | --- | --- |
| High- Low | 2.45 | 2.56 | 2 | 0 | 9 | 0.55 | -0.058 |
| Low-Recovered | 3.22 | 3.67 | 3 | 0 | 11 | 0.60 | 0.19 |

**Figure 5.**
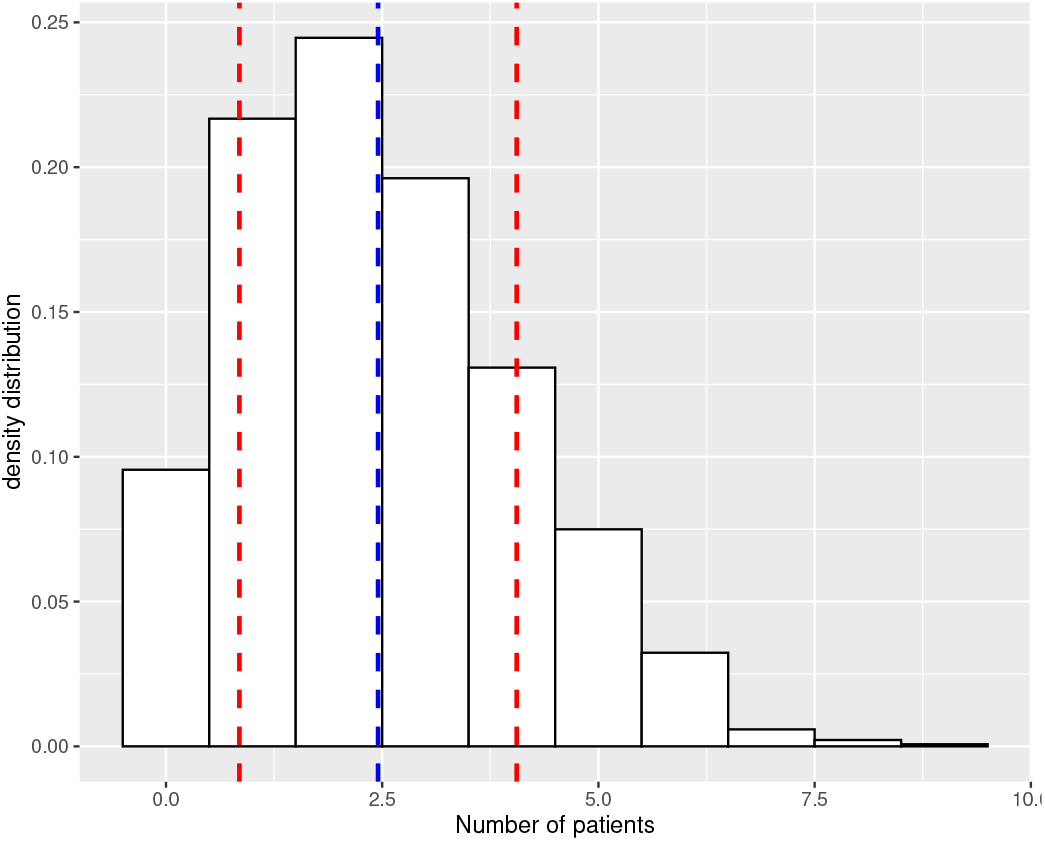
Daily distribution of the number of patients that move from high-acuity to low-acuity. (Blue line represents the mean and the red lines are the one standard deviations from the mean.)

**Figure 6.**
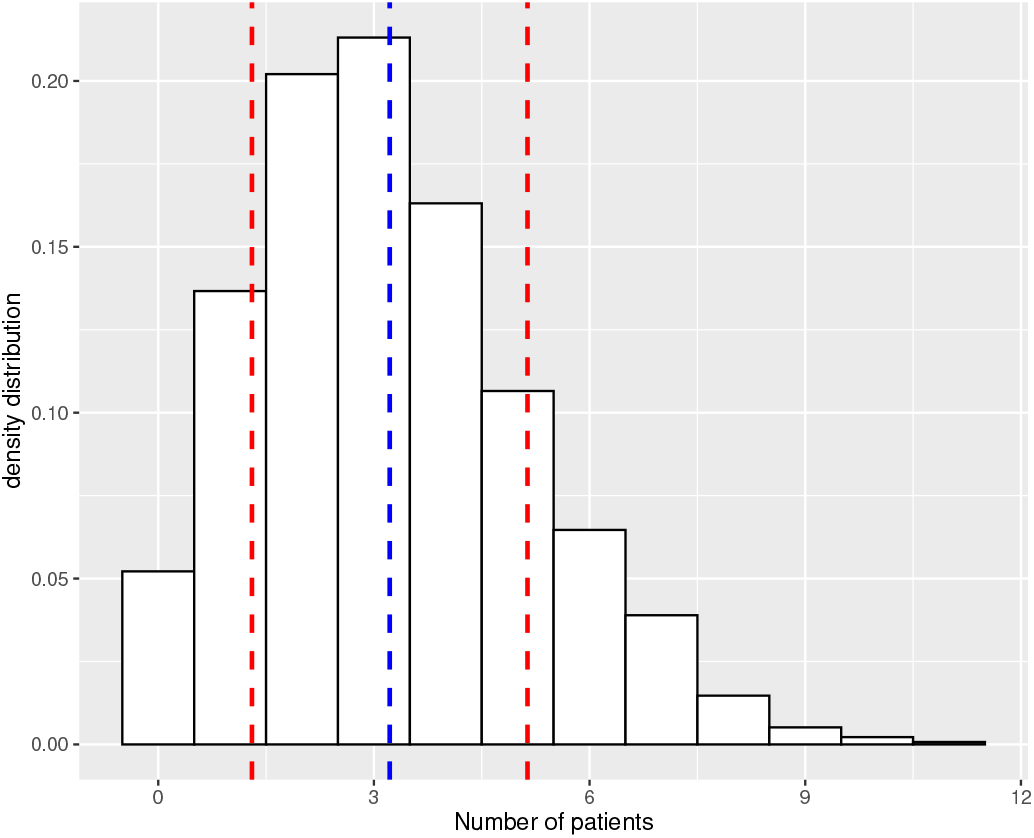
Daily distribution of the number of patients that move from low-acuity to recovered. (Blue line represents the mean and the red lines are the one standard deviations from the mean.)

On average, the daily transition matrix from one acuity level to another is given in Table 4.

**Table 4.** Average daily transition probability

|  | High | Low | Recovered | Discharged |
| --- | --- | --- | --- | --- |
| High | 0.7760 | 0.1360 | 0.0020 | 0.0690 |
| Low | 0.1505 | 0.4342 | 0.0040 | 0.346 |
| Recovered | 0.0700 | 0.2250 | 0.0899 | 0.6140 |
| Discharged | 0 | 0 | 0 | 1 |

## 4. Methodology

Patient flow through the SDU in most hospitals is assumed to come from two areas: either directly from the ICU, or as a direct entry if the ICU is full. Patients are admitted to the SDU directly from the ICU once they are deemed to be less acute for the ICU, but not so sick that they require ICU care. Alternatively, acute patients who cannot be admitted to the ICU due to congestion are admitted to the SDU. A direct entry into the SDU is another example of an Alternative Level of Care (ALC). In other words, if the ICU is too busy to accept a patient right away, patients may occasionally be admitted to the SDU before receiving ICU treatment. For instance, certain SDUs, as described in Cady et al. (1995); Eachempati et al. (2004), exclusively admit post-ICU patients, whereas other SDUs permit various admission patterns. This study used the same assumption as Cady et al. (1995) that all SDU patients have left the ICU.

Patients arriving at the ICU are assumed to be high-acuity patients only and are admitted immediately to the ICU if there is a bed. In reality, some patients admitted to the ICU may have a NEMS score less than 25. However, due to the presence of multiple factors described in Table 2, they are treated in the ICU. This study assumes only two types of patients in the ICU: high and low-acuity. High-acuity patients have NEMS scores greater than or equal to 25 and the low-acuity patients have NEMS scores less than 25 (Rodrigues et al. 2018). At a decision epoch, which is continuous, a patient flow decision must be made. When the ICU is full and a high-acuity patient arrives at the ICU, he or she may be rejected, or a less acute patient may be stepped down to make a space, or a high-acuity patient may be prematurely stepped down. A critical patient who is admitted to the ICU will be treated until either reaching a stable enough low-acuity state or stepping down to the SDU.

This study proposes a Markov decision process (MDP) to model patient flow dynamics. The objective is to maximize “the net health service benefit” of the system’s flow under a congested environment. The accumulation of rewards and costs associated with each of the actions under a policy gives the net health service benefit of a policy. The goal is then to find the set of actions that maximizes the net health service benefit.

### 4.1. State Space and Action Set

At a decision epoch, *t* ∈ [0, ∞) considered to be continuous, the decision-maker has a set of decisions to make: (i) Admit or reject an arriving patient, if any, to ICU (Rejection can be to an alternative level of care or an off-site service), (ii) step-down to the SDU or retain a low-acuity patient, (iii) prematurely discharge from the SDU to the ward or retain a recovering patient (iv) prematurely step down from the ICU to the SDU or retain a high-acuity patient (If the ICU is full and there is an arrival, premature step-down may be considered). Because this study looks at decisions under congestion, the system state is defined in terms of the congestion zone or the last-bed zone (Azcarate et al. 2020). The system state is denoted by 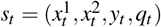 where 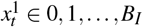 is the number of high-acuity patients occupying the *B*_*I*_ congestion zone of the ICU, 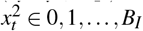 is the number of low-acuity patients occupying the *B*_*I*_ congestion zone of the ICU, *y*_*t*_ ∈ 0, 1, …, *B*_*S*_ is the number of low-acuity patients occupying the *B*_*S*_ congestion zone of the SDU, and *q*_*t*_ ∈ 0, 1 is the number of arriving patients.The state-space at time epoch *t* is defined as:

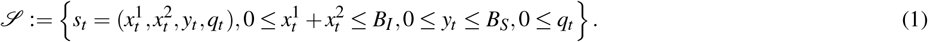

For each state *s*_*t*_ ∈ 𝒮, *z*_*t*_ = (*z*_1*t*_, *z*_2*t*_, *z*_3*t*_, *z*_4*t*_), 𝒜 (*s*_*t*_) denote a feasible action that can be taken. The action state is given as:

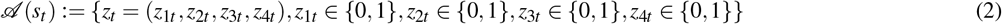

where *z*_1*t*_ = 1 denotes admission otherwise rejection; *z*_2*t*_ = denotes step-down to the SDU, otherwise, retention; *z*_3*t*_ = 1 denotes discharge to the ward, otherwise retention; and, 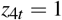 denotes premature step down otherwise retention. Every action must satisfy the capacity constraint, expressed as

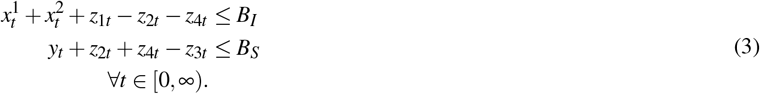

These actions are taken so that the system cannot accept more than its capacity. The rejection of arriving patients when the capacity is exceeded guarantees that.

If there are enough beds in the ICU, then the decision may seem simple: admit all arriving patients. If there is enough capacity in the SDU, then all low-acuity patients in the ICU can step down. If the ICU is not full, there is no need to discharge anyone prematurely (Li et al. 2018). The main concern that motivates this research is the problem of congestion, or “the last bed” in both the ICU and the SDU. Rejecting patients is undesirable and may be impractical. If congestion exists in the ICU, patients that would have been admitted may wait in other hospital units (e.g. ED, surgical wards, general wards) but maybe recorded as ICU patients. Keeping a patient in the ICU when that patient is supposed to be discharged to the SDU due to SDU congestion is also undesirable because since this patient consumes resources that he or she no longer needs and prevents others from using these resources. It may also not be in the hospital’s best interest to have too many idle beds. Because this study considers only congestion, instead of using the whole the whole system capacity of the units to build the state space, only the congestion zone is considered. For simplicity, the last two beds of the ICU and the last bed of the SDU corresponding to the congestion zone is considered. The problem becomes the last bed problem in the medical literature (Donaldson et al. 2000; Rodziewicz et al. 2018; Azcarate et al. 2020).

Table 5 describes the state space. The first policy allows admit or reject, step-down or retain a low-acuity patient in the ICU, and premature discharge or not from the SDU. The actions are coded as tabulated in Table 6. A feasible combination of these actions is described in table 7. The second policy allows in addition to the actions of the first policy, premature step-down of high-acuity patients. The actions are coded as shown in Table 8. A feasible combination of these values gives a complete description of the action space for the first case as shown in Table 9.

**Table 5.**
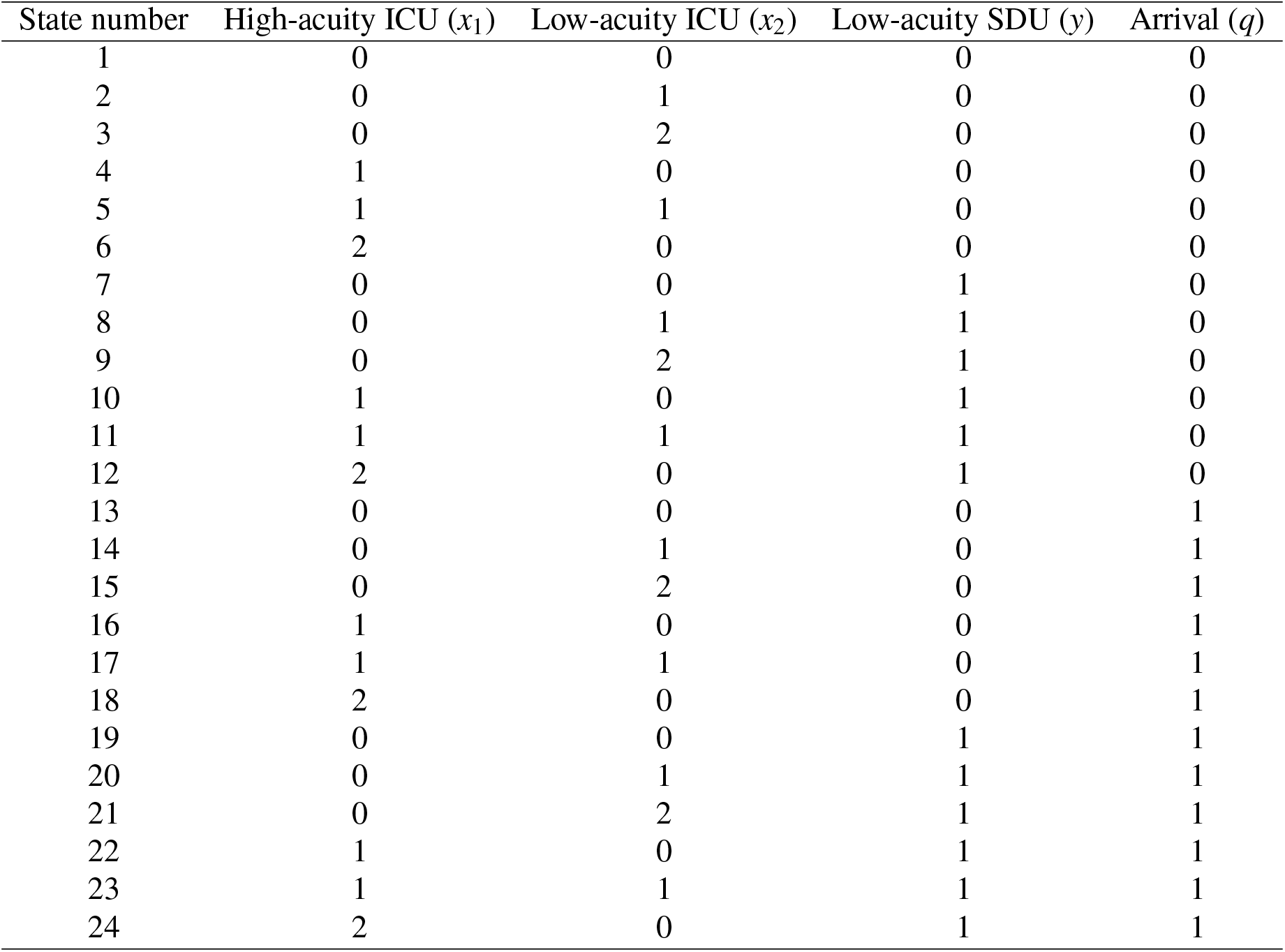
Description of the state space of the solved system

**Table 6.** First policy actions

| Action | Value | Description |
| --- | --- | --- |
| a1 | 0,1 | Admission of high-acuity to ICU |
| a2 | 0,1 | Step-down of low-acuity to SDU |
| a3 | 0,1 | Discharge of low-acuity out of SDU |

**Table 7.**
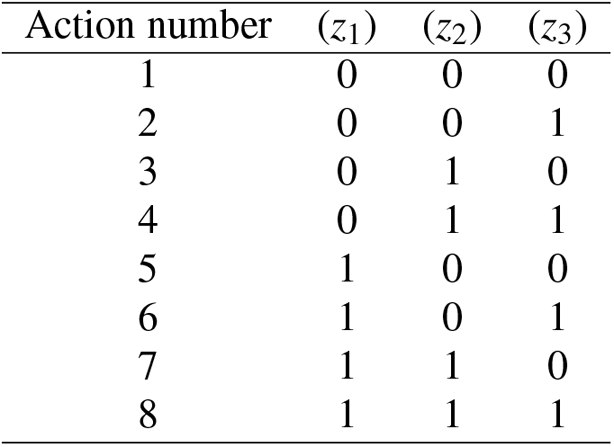
Feasible actions under the first policy

**Table 8.** Second policy actions

| Action | space | Description |
| --- | --- | --- |
| a1 | 0,1 | Admission of high-acuity patients to ICU |
| a2 | 0,1 | Step-down of low-acuity patients to SDU |
| a3 | 0,1 | Discharge of low-acuity patients out of ICU |
| a4 | 0,1 | Premature Step-down of high-acuity patients |

**Table 9.**
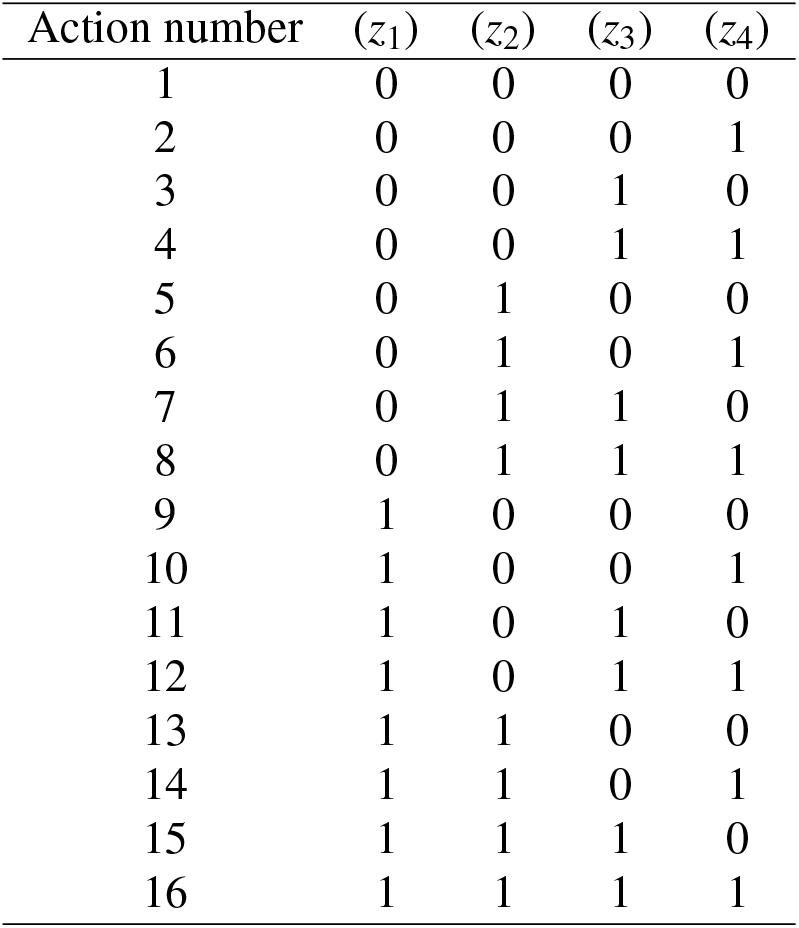
Feasible actions under the second policy

| Action number | ($z_1$) | ($z_2$) | ($z_3$) | ($z_4$) |
| --- | --- | --- | --- | --- |
| 1 | 0 | 0 | 0 | 0 |
| 2 | 0 | 0 | 0 | 1 |
| 3 | 0 | 0 | 1 | 0 |
| 4 | 0 | 0 | 1 | 1 |
| 5 | 0 | 1 | 0 | 0 |
| 6 | 0 | 1 | 0 | 1 |
| 7 | 0 | 1 | 1 | 0 |
| 8 | 0 | 1 | 1 | 1 |
| 9 | 1 | 0 | 0 | 0 |
| 10 | 1 | 0 | 0 | 1 |
| 11 | 1 | 0 | 1 | 0 |
| 12 | 1 | 0 | 1 | 1 |
| 13 | 1 | 1 | 0 | 0 |
| 14 | 1 | 1 | 0 | 1 |
| 15 | 1 | 1 | 1 | 0 |
| 16 | 1 | 1 | 1 | 1 |

### 4.2. Health service benefit Rewards and Costs

The reward and cost structure of this study is defined as the health service benefit of an individual in the hospital. Because ICU managers desire to serve the high-acuity patients in the ICU and the low-acuity patients in the SDU, the reward comes from two sources: admitting patients to the ICU is given a higher reward, and stepping down a low-acuity patient from the ICU to the SDU to accommodate high-acuity patients in the ICU is rewarded. A reward, *r*_*h*_, is associated with every admission and a reward, *r*_*l*_, is associated with every natural step-down. Natural discharges are not rewarded not to double reward. The reward, *r*_*l*_, is viewed as the profit of not using the ICU with higher cost and using the SDU with lesser cost, but with the same result. It is considered to be the difference between the cost of rejecting an arriving patient to the ICU and the cost of the current patient using the ICU for a full recovery. The undesirable events are then seen as elements that contribute to cost. Rejecting patients, premature step-down, keeping a low-acuity patient in the ICU and premature discharge of patient that has not fully recovered all contribute to the cost of health service benefit to individual patients in the hospital. A cost, *c*_*h*_, is associated with every high-acuity patient rejected; a cost, *c*_*l*_, is associated with every overstay of a low-acuity patient in the ICU; a cost, *c*_*d*_, is associated with every low-acuity patient prematurely discharged out of the SDU; and a cost, *c*_*p*_, is associated with a premature step-down out of the ICU. The cost of a premature step down represents the cost of incomplete service in the ICU. The reward associated with the natural step down is considered the difference between the cost of rejecting an arriving patient at the ICU and the cost of the current patient using the ICU for a full recovery. The cost of a premature step down represents the cost of incomplete service at the ICU.

Health service benefit rewards are gains and health service benefit costs are losses. The accumulation of rewards and costs defined above produces the net health service benefit. This research aims to obtain the decision structure to maximize the discounted net health service benefit over one quarter. The structure of the net health service benefit can be written as:

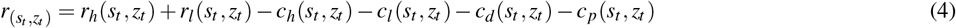

It is assumed that the cost of ICU refusal is equal in absolute terms to the reward for ICU use. Given that the SDU is poorly equipped and less monitored, it was assumed that its absolute service cost was less than that of the ICU. Rejecting a patient has a greater negative effect than prematurely stepping down a patient. Likewise, premature step-down has a greater negative effect than premature discharge from the SDU. For the toy example, the baseline values for the computation are set as follows: the reward for admitting a patient is set to 100, the reward for stepping down a patient is 25, the cost for rejecting a patient is 100, the cost for overstay in the ICU is 50, the cost for premature discharge from the SDU is 25, and the cost for premature step-down of a high-acuity patient is set to 80. These values are chosen as relative weights of the consequences of each action. Measuring the effect of these actions is difficult because such research would be unethical. The idea is to start with a naive relative weight for each action. In this way, sensitivity analysis of these values for its robustness was performed.

### 4.3. Value Function and Transition Probability

The value function estimates how good it is for the decision-maker to perform a given map of actions to the state. Every policy is measured by a value function of each policy. A policy *π* is a distribution of a set of the feasible action in each state (a mapping from each state, *s* ∈ 𝒮, and action, *z* ∈ 𝒜, to the probability *p*_(*s,z*)_ of taking an action *z* when in state s). In other words, it is a mixed policy. The value function in state *s* under a policy *π* at time *t*, denoted as *V*_*π*_ (*s*_*t*_), give the expected long run value of the discounted rewards when starting in *s* and following the policy *π* thereafter. It is defined by

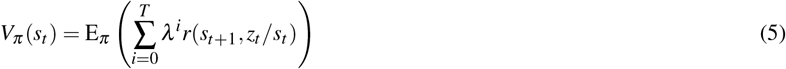

where *V*_*π*_ (*s*_*t*+1_*/s*_*t*_) is the reward function at time *t* + 1 given that action *z* is taken in state *s* at time *t* and the time horizon, *T* = ∞, is an infinite time horizon. The infinite horizon was considered because decision epochs happen continuously, whereas arrivals occur randomly. The optimal value function of the MDP model specifies the maximum expected reward over the infinite horizon for each state and satisfies Bellman’s optimality equation (Puterman 2014) for all *s*_*t*_ ∈ **S** defined by

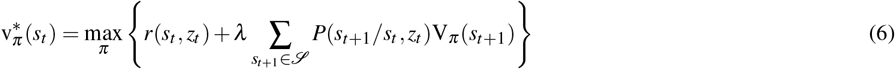

The objective is to determine the optimal policy *π*\* of the MDP. This policy specifies the distribution of the actions that optimize the value function for each state and is given by

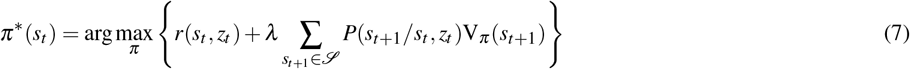

*P*(*s*_*t*+1_*/s*_*t*_, *z*_*t*_) is the probability of transiting from state *s* to another when action *z* is taken at time *t*. The transition probability describes the interactive combination of the progression of a patient’s health status from one acuity level to another, the random arrivals and the actions taken and is given by

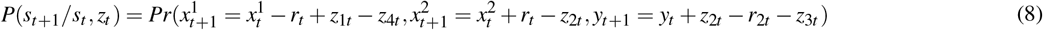

where 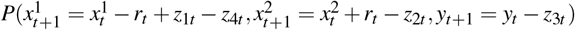 is the probability that at time *t* + 1, 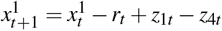 is the number of acute patients in the ICU, 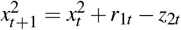 is the number of recovering patients in the ICU, and *y*_*t*+1_ = *y*_*t*_ + *z*_2*t* −_ *r*_2*t* −_ *z*_3*t*_ is the number of patients in the SDU, where *r*_*t*_ is the number of people who recover from the *x*_*t*_, and *r*_2*t*_ is the number of natural recoveries who left the SDU. At time epoch *t*, the system state is 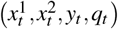. Between time epoch *t* and *t* + 1, three processes are considered to influence the state transition. Arrivals *q*_*t*_, recovery of 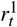 high-acuity patients to low-acuity patients in the ICU, and complete recovery of 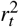 low-acuity patients that move out of the system at time *t* + 1. The system state at time *t* + 1 becomes 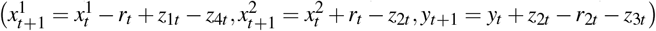. This is depicted in the diagram below.

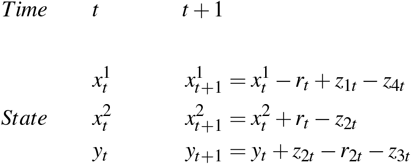

The transition probability is obtained from three random events: arrival, recovery in the ICU, and recovery in the SDU. *q*_*t*_ arrivals occur at time *t* with probability *Pr*(*q*_*t*_). This random arrival is independent of patients’ recovery rates and of hospital management. *r*_1*t*_ recovered from high-acuity to low-acuity with probability *Pr*(*r*_1*t*_) and *r*_2*t*_ recovered from low-acuity to recovered with probability 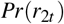. Because these three processes are independent, the transition probability can be safely approximated as:

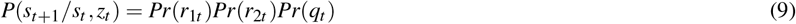

and the distribution of each of these events is estimated from data, as shown in Section 3 for the simulation. Because a Poisson distribution is the limit of a binomial distribution with parameter *p* = *λ/n*, where *λ* is the Poisson rate, and n, the number of trials, approaches infinity. Because this study considers continuous-time epochs, it can be assumed that at most one patient may arrive, at most one patient may recovery from high-acuity to low, at most one patient may recover from low-acuity to recovered, and at most one patient will arrive. Each process can then be a Bernoulli process with parameter *p*. Because the assumption on *n* is subjective, it was assumed that an event occurring or not was equi-probable. Hence, for the computation, the transition probability from state *s*_*t*_ to state *s*_*t*+1_ is 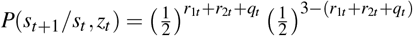 with *r*_1*t*_, *r*_2*t*_, *q*_*t* ∈_ {0, 1}.

*z*_1*t*_ is admitted at time *t, z*_2*t*_ is stepped down to the SDU at time *t, z*_3*t*_ is prematurely discharged at time *t*, and *z*_4*t*_ is prematurely stepped down from the ICU at time *t*. The objective is to determine the distribution of the joint probability mass function of these actions and use it to the action with the maximum probability weight as the approximate optimal action in various states.

### 4.4. Solution Methodology

Several methods have evolved for solving MDPs and dynamic processes in general. Solution methods for infinite-horizon problems use policy iteration, value iteration and linear programming, whereas finite-horizon problems are mostly solved using backwards induction algorithms. This paper uses the linear programming method developed by Schweitzer and Seidmann (1985) with more recent expansions by (Adelman 2007; De Farias and Van Roy 2003; Puterman 2014) to solve the MDP due to its simplicity and easy reproducibility. Bertsekas and Tsitsiklis (1995); Powell (2007); Manne (1960); Adelman (2007); De Farias and Van Roy (2003); Puterman (2014) provide an extensive literature on linear programming methods for solving MDPs. From Powell (2007) and Puterman (2014), it is known that, if 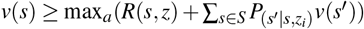 where 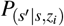 is the transition probability, then *v*(*s*) is an upper bound on the value of being in each state. This means that the optimal value function can be obtained and the optimal actions determined through backward induction by solving the following linear program

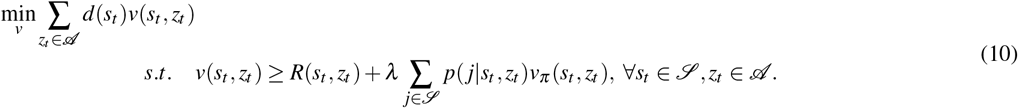

where *d*(*s*_*t*_) is any positive value. Alternatively, the solution of the dual of Equation 10 shown in Equation 11 provides the distribution of the actions in each state (Denardo 2012).

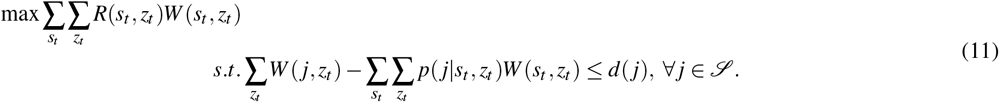

where the normalized *W* (*s*_*t*_, *z*_*t*_), (*s*_*t*_ ∈ *S, z*_*t*_ ∈ 𝒵) are interpreted as the steady-state probabilities that action *z*_*t*_ is applied when the system visit state *s*_*t*_ at the typical transition. There are in total # 𝒮 constraints, where # 𝒮 represents the total number of states in the states space. The cost function 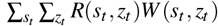 represents the steady-state average reward per transition. From Ross (2014) and Wang et al. (2007) by strong duality, the optimal objective value of the dual LP equals the optimal objective value of the primal LP. Therefore, given a solution to the dual, the optimal action can be approximated directly by a much simpler transformation as

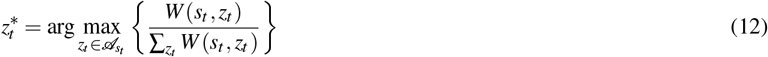

where 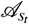 is the set of all the actions possible in state *s*_*t*_.

## 5. Results

### 5.1. Optimization Results

Equation 11 and the probabilities of each of the actions in every state were computed. The action with the maximum probability in the state is the optimal action. Programming was done in R using the lpSolveAPI package, a 64 bit laptop with 32 GB memory and an Intel(r) Core i7-4600U CPU at 2.69 GHz.

The optimization and the computation were done as described in sections 3.5 and 4.1. Table 10 presents the optimal actions in each state for Policy 1. In states without arrival, the possible actions are reduced to step-down or retain low-acuity patients in the ICU to the SDU and/or discharge or retain a low-acuity patient in the SDU. Discharges occur only when there is at least one low-acuity patient in both the ICU and the SDU, with or without arrivals. Every discharge out of the SDU has been triggered by a step-down from the ICU, and the presence of two low-acuity patients in the system. Either the two low-acuity patients are in the ICU or there is one in the ICU and one in the SDU. Whenever there is a space in the SDU and there is a low-acuity patient in the ICU, a step-down is triggered. Once an arrival takes placeif there is a low-acuity patient in the ICU, a step-down is triggered. Whenever there is an arrival and the ICU has an empty bed, the person is admitted. In the state where two ICU beds are taken by low-acuity patients, the SDU is empty, and an arrival takes place, the recommended action is to admit and step down. In general, accept arrivals when you can, step down when you can, and discharge when needed.

**Table 10.** Case 1 Comparative Optimal Decision in States with and without Arrival

| State | High<br>acuity<br>ICU ($x_1$) | Low<br>acuity<br>ICU ($x_2$) | Low<br>acuity<br>SDU ($y$) | Action taken without arrival | Action taken with arrival |
| --- | --- | --- | --- | --- | --- |
| 1 / 13 | 0 | 0 | 0 | admit 0, step-down 0 and discharge 0 | admit 1, step-down 0 and discharge 0 |
| 2 / 14 | 0 | 1 | 0 | admit 0, step-down 1 and discharge 0 | admit 1, step-down 1 and discharge 0 |
| 3 / 15 | 0 | 2 | 0 | admit 0, step-down 1 and discharge 0 | admit 1, step-down 1 and discharge 0 |
| 4 / 16 | 1 | 0 | 0 | admit 0, step-down 0 and discharge 0 | admit 1, step-down 0 and discharge 0 |
| 5 / 17 | 1 | 1 | 0 | admit 0, step-down 1 and discharge 0 | admit 0, step-down 1 and discharge 0 |
| 6 / 18 | 2 | 0 | 0 | admit 0, step-down 0 and discharge 0 | admit 0, step-down 0 and discharge 0 |
| 7 / 19 | 0 | 0 | 1 | admit 0, step-down 0 and discharge 0 | admit 1, step-down 0 and discharge 0 |
| 8 / 20 | 0 | 1 | 1 | admit 0, step-down 1 and discharge 1 | admit 1, step-down 1 and discharge 1 |
| 9 / 21 | 0 | 2 | 1 | admit 0, step-down 1 and discharge 1 | admit 1, step-down 1 and discharge 1 |
| 10 / 22 | 1 | 0 | 1 | admit 0, step-down 0 and discharge 0 | admit 1, step-down 0 and discharge 0 |
| 11 / 23 | 1 | 1 | 1 | admit 0, step-down 1 and discharge 1 | admit 1, step-down 1 and discharge 1 |
| 12 / 24 | 2 | 0 | 1 | admit 0, step-down 0 and discharge 0 | admit 0, step-down 0 and discharge 0 |

Table 11 describes the optimal actions selected in the various states under Policy 2. In states where there is no arrival, the possible actions are reduced to step-down low-acuity patients from the ICU to the SDU and/or discharge a low-acuity patient from the SDU and/or premature discharge of a high-acuity patient. It can be observed that as long as there is no arrival, no premature step-down is necessary. Note that the only state in which a premature step down is allowed is state 18, i.e. when all ICU beds are occupied, one SDU bed is available and an arrival takes place. In the case one of the ICU patients is prematurely stepped down and the arriving high-acuity patient admitted.

**Table 11.**
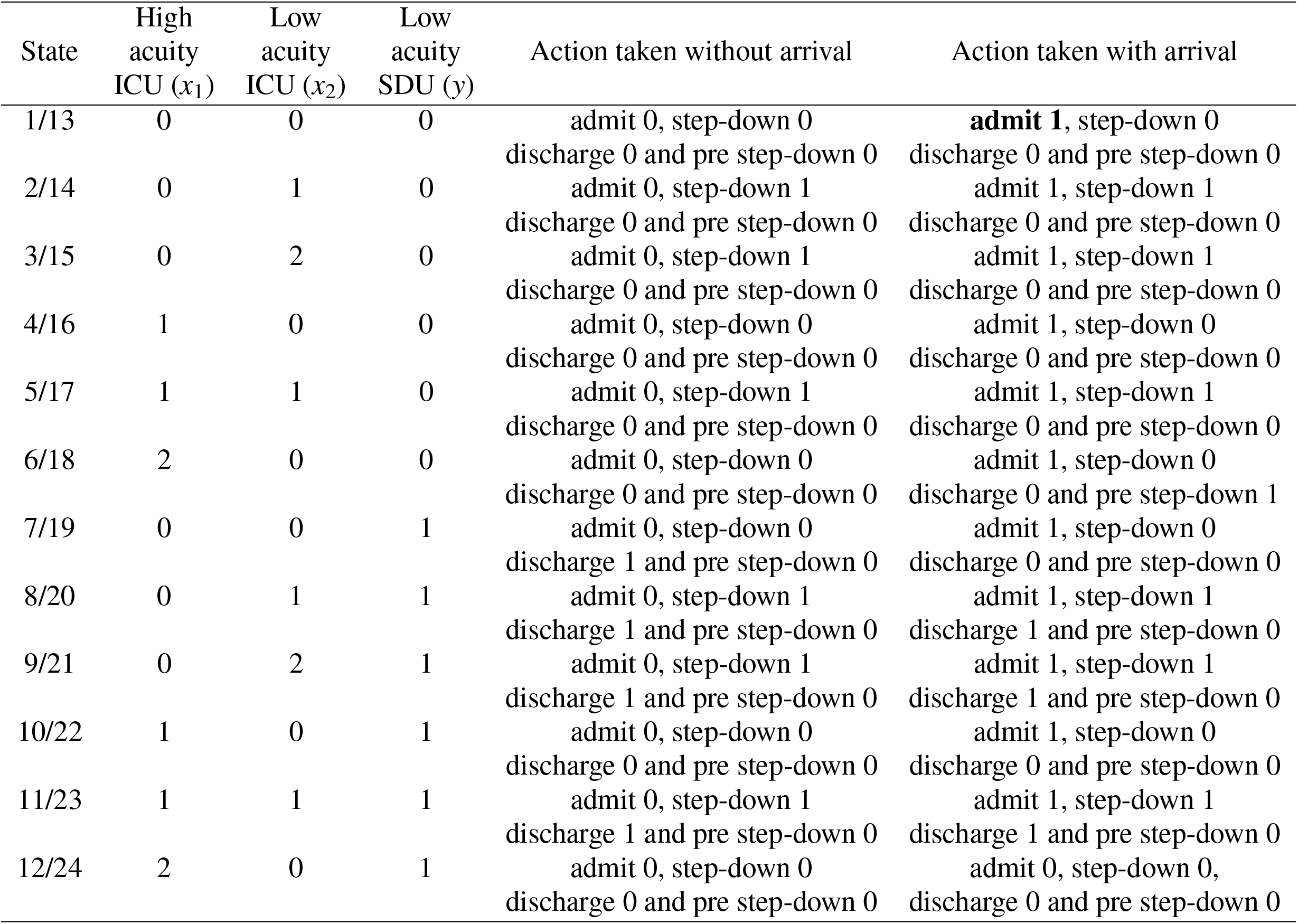
Case 2 Comparing the optimal decision in states with and without Arrival

In general, a discharge is triggered by a step-down of a low-acuity patient in the ICU, whenever there are at least two low-acuity patients in the system and a low-acuity patient is occupying an SDU bed. The only time admission is denied is when the ICU is full and the SDU is also full. If all the patients in the ICU are high-acuity patients, there may be high-acuity patient in the SDU as well. In other words, the patient occupying the last bed in the SDU may also be a high-acuity patient. In general, it is better to allow the recovering patient in the SDU to recover than to discharge him, with a cost, and admit an arriving patient in the SDU with another cost.

### 5.2. Sensitivity analysis of costs and rewards

A sensitivity analysis was undertaken to check how changes in the baseline costs and rewards change the optimal action in the different states. The base cost/reward parameters used in the model are as follows. The reward for admitting a patient is 100, the reward for stepping down a patient is 25, the cost for rejecting a patient is 100, the cost for overstay in the ICU is 50, the cost for prematurely discharging a patient is 25 and the cost of premature step-down of a patient is 80. In general, actions are robust to rewards and costs associated with each action within the neighbourhood. A large variation in the cost and reward parameters is necessary for a fundamental change in the decision. For example in state (0,2,0), i.e. 0 high-acuity in the ICU, 2 low-acuity in the ICU and zero low-acuity in the SDU, using the first policy, rewards must be increased from 100 to 800 before a change in the action from action (0,1,0) occurs i.e., admit 0, step-down 1, and discharge 0 to action (0,0,0), admit 0, step-down 0 and discharge 0 (see Table 12 first row).

**Table 12.**
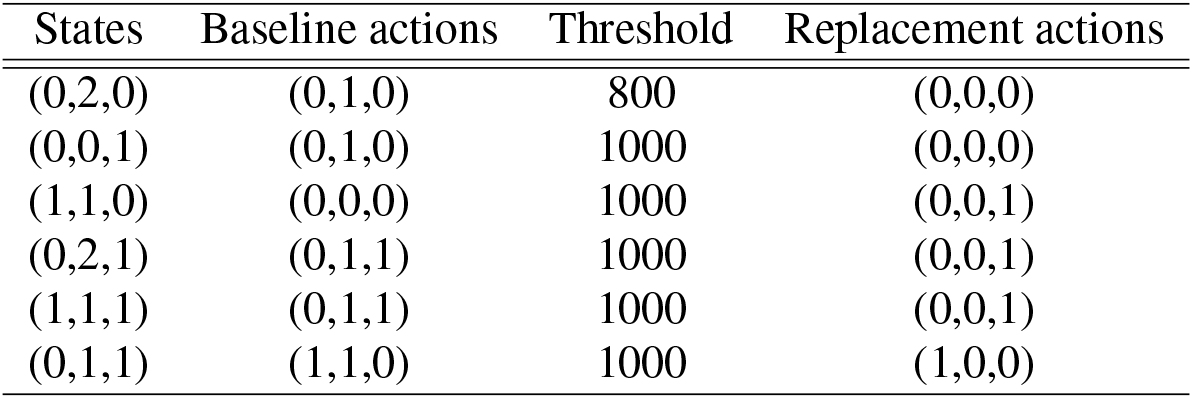
Sensitivity Summary of Model without Premature Step-down when admission reward and rejection cost are increased. The replacement actions are used when the value is greater than the threshold.

The first policy, assumes that the step down reward and the cost of overstay is always less than the rejection cost. With that assumption, the model is always robust to step down reward and the cost of overstay. Only the admission reward or rejection cost have the same change as summarized in Table 12. Note that the Tables summarize only states that show variation. When the system is in state (1,1,0), i.e., one high-acuity, one low-acuity in the last ICU bed and no patient in the last SDU bed, the baseline action is to do nothing, but when the admission reward is increased from 100 to 1000, the replacement action becomes stepping down the low-acuity patient from the ICU to the SDU. When the admission reward or the rejection cost increases, the system tends to perform fewer step-down actions. In the second policy, increasing discharge cost while keeping all other rewards and cost constant significantly affects decisions in only two states. In states (0,2,1,1) and (1,1,1,1), action (0,1,1,1) is replaced by action (0,0,0,0). In other words, when there are at least two low-acuity patients in the system, and the discharge cost is high, the optimal action recommended is to do nothing. Table 13 summarizes the variations observed when the reward for admission, the cost for rejection, and the step-down reward are increased. Increasing the cost of overstay while keeping all other rewards and costs constant affects two states. In state (1,0,0,0), the decision changed from action (0,0,0,0) to action (0,0,0,1). In state (1,0,0,0), the decision changed from action (0,0,1,0) to action (0,0,0,0). In state (1,0,0,1), the decision changed from action (1,0,0,0) to action (0,0,0,0). When the ICU is full and the SDU is full, no matter how much the cost is decreased, premature step-down is never a better option (See Table 9 for actions). In state (2,0,0,1), when the cost has increased, it is not a better option to perform a premature step-down. In state (1,0,0,0), when the cost is low, the system can afford to perform a premature step-down and admit once there is a space in the SDU. In state (1,0,0,0) even though the ICU is not full, when the cost of premature step-down is low, the step down is recommended.

**Table 13.** Sensitivity Summary of Model with Premature Step-down.(AR: Admission Reward, RC: Rejection Cost, SR: Step-down Reward)

| Parameters | States | Baseline action | Replacement action | Threshold |
| --- | --- | --- | --- | --- |
| AR | (1,0,0,0) | (0,0,0,0) | (1,0,0,0) | 250 |
|  | (2,0,0,0) | (0,0,0,0) | (1,0,0,0) | 500 |
|  | (1,0,1,0) | (0,0,0,0) | (0,0,0,1) | 1000 |
|  | (1,0,0,1) | (0,1,0,0) | (1,0,1,0) | 1500 |
|  | (0,0,1,1) | (0,1,0,0) | (0,1,0,1) | 500 |
| SR | (0,0,1,0) | (0,0,0,1) | (0,0,0,0) | 1000 |
|  | (2,0,1,0) | (0,0,0,0) | (1,0,0,1) | 1000 |
| RC | (2,0,0,0) | (0,0,0,0) | (1,0,1,0) | 1000 |
|  | (0,2,1,0) | (0,0,1,1) | (0,0,0,0) | 1000 |
|  | (1,1,1,0) | (0,0,1,1) | (0,0,0,0) | 1000 |
|  | (1,0,0,1) | (0,1,0,0) | (1,0,1,0) | 2000 |
|  | (0,1,1,1) | (0,1,1,1) | (0,1,0,0) | 2000 |
|  | (2,0,1,1) | (0,0,0,0) | (0,0,0,1) | 2000 |

### 5.3. Simulation

The model was built using Simul8 6.0 (See Figure 8). The software was chosen for its availability, flexible coding, simplicity, and interactive display of sequential events. In the simulation frame, there were 30 ICU beds and 12 SDU beds. Decision epochs were continuous. The optimal decisions were triggered only when the system was in the congestion zone. Arrivals followed a Poisson distribution with a rate of 6.3 patients/day (Approximation from the hospital data, see Figure 4). As estimated in Section 3, the recovery process from high to low acuity followed a Poisson distribution with a rate of 2.45 patients/day, recovery from low-acuity to recovered was a Poisson process with a rate of 3.22 patients/day, the discharge process from the system to elsewhere also followed a Poisson distribution with a rate of 4.47 patients/day, and the death process was also Poisson with a rate of 1.24 patients/day. Decision-making was a continuous process triggered by any of the previous processes. When there was space in the ICU, an arriving patient was automatically admitted. The internal decisions in the ICU and SDU were coded into the system as tabulated in Table 10 or Table 11 depending on the policy considered. In the ICU, the low-acuity patients were then distinguished from the high-acuity patients. If there was space in the SDU, the low-acuity patients were moved to the SDU.

**Figure 7.**
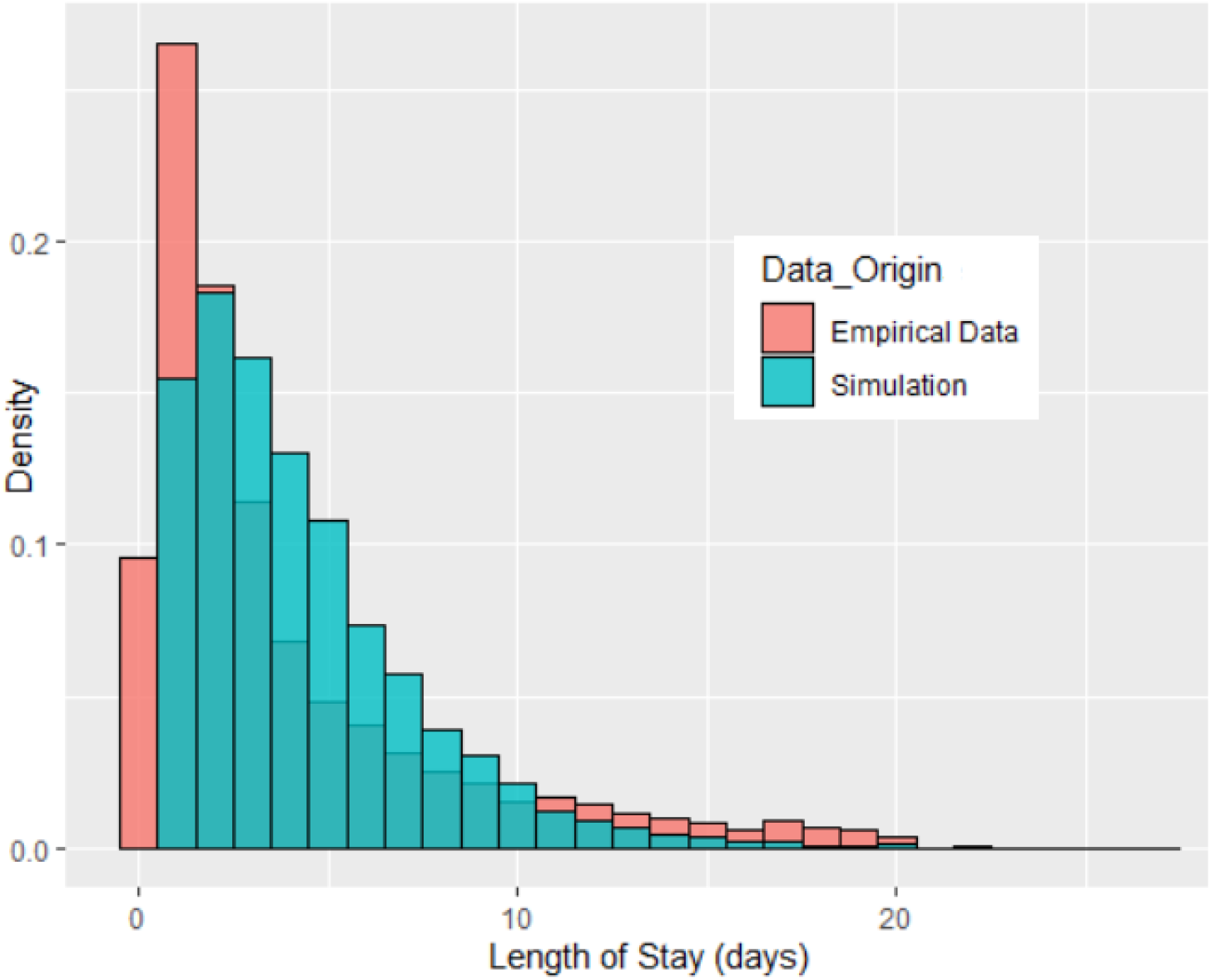
Length of Stay Distribution

**Figure 8.**
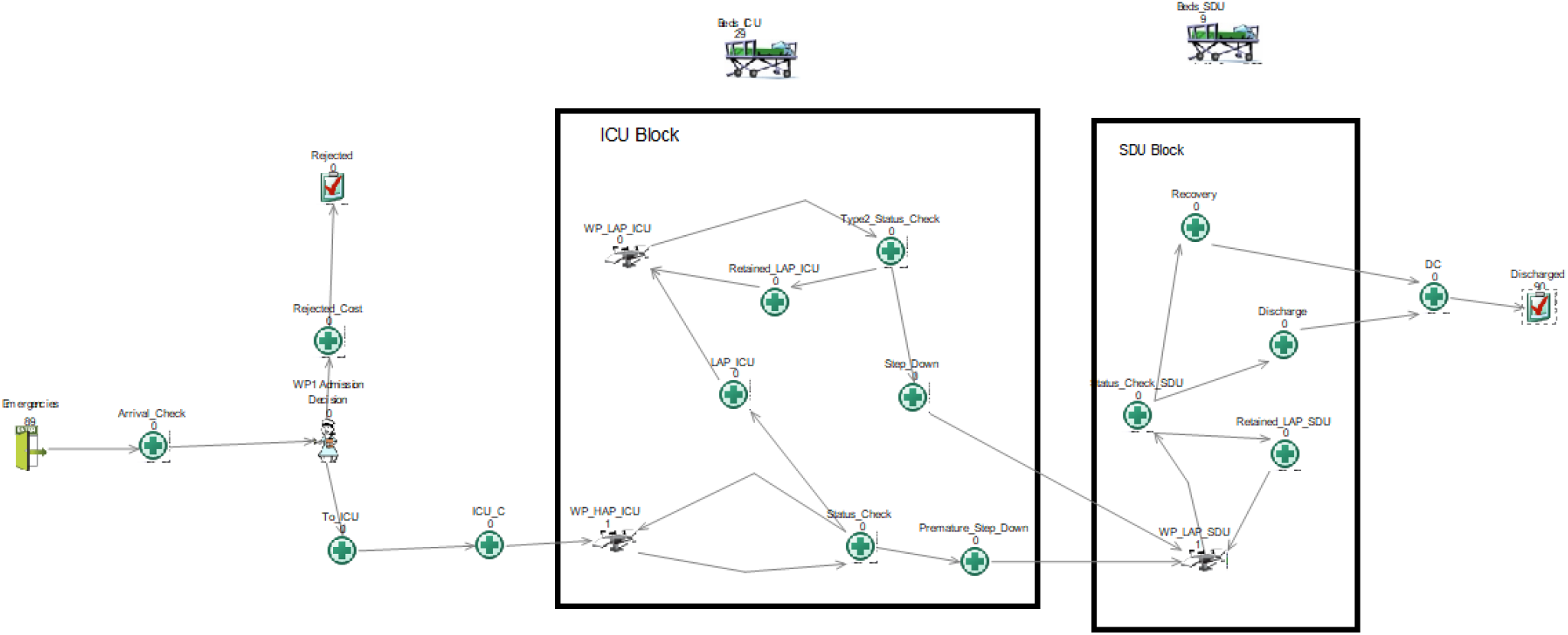
Screenshot of Simulation in Simul8

Two methods were used to validate the model: experts in-person face validation, and the comparison of parameter estimates and simulation results without implementation of the optimal decisions. The average throughput was within 5.78% of the empirical estimate. Total LOS was lower but with a standard deviation of 3.4, resulting in no statistically significant difference compared to the empirical data. The mean LOS was found to be within 9.27% of the empirical value. 7 shows a plot of the superimposed distributions of the empirical and simulated LOS. Given that the empirical data included some special patients who had been in the ICU for months, those observations were removed to obtain statistical significance.

Each policy was run for ten months with 300 replications of 50000 trials. Only records of the last four months of the simulation were reported to obtain stable results. The number of replications was recommended by the replication calculator, which is a a function embedded in Simul8. Precision was set to a 95% confidence interval. A different random seed was used for each of the runs. The costs incurred, the number of patients rejected and the number of patients prematurely stepped down were examined under the two scenarios to evaluate and compare the performance of the decision policies.

### 5.4. Simulation Results

The estimated performance indicators of the system when the arrival rate was six patients per day (from empirical data) are tabulated in Tables 14 and 16. From Table 14, the results suggest that on average, no patient was rejected with this average arrival rate. Policy 2 prematurely stepped down on average about 47% (253) of the patients admitted causing an average overstay of 10% (55) patients. Conversely, Policy 1 overstayed only 2.57 % (13) patients. ICU bed utilization was moderate, but SDU bed utilization was high as observed in real life. Policy 2 led on average to a lesser utilization of the ICU (35%) compared to Policy 1 (49%). Policy 1 gave a less congested SDU utilization of about 58% on average compared to 71.8% under Policy 2. In terms of health service benefit and cost, Policy 1 had an average cost per admission of 13.57, and, an average total reward of 125.24, leading to a net benefit of 111.67 per patient admitted. In contrast, Policy 2 had an average cost per admission of 55.40, and average total reward of 113.28, leading to a net average benefit of 57.896 per patient admitted.

**Table 14.** Comparative Patients Performance Measures

| Indicator | Low 95 % | Simulation Average | Up 95% | Emp data Estimate | Difference |
| --- | --- | --- | --- | --- | --- |
| Throughput (Patients/year) | 1332 | 1366 | 1399 | 1287 | -5.78% |
| Average LOS (days) | 4.04 | 4.11 | 4.18 | 4.53 | -9.27% |
| Standard Deviation (days) | 3.354 | 3.4 | 3.447 | 4.13 | -17.67% |

**Table 15.** Patients flow performance measures over four months.

| Performance measures | Policy 1 |  |  | Policy 2 |  |  | diff |
| --- | --- | --- | --- | --- | --- | --- | --- |
|  | Low 95% | Av | Up 95% | Low 95% | Av Total | Up 95% |  |
| Admitted | 462 | 515 | 568 | 534 | 537 | 540 | -22 |
| Rejected | 0 | 0 | 0 | 0 | 0 | 0 | 0 |
| Pre-stepped down | 0 | 0 | 0 | 251.46 | 253.22 | 254.97 | -253.22 |
| ICU overstay | 4.79 | 13.25 | 21.71 | 52.43 | 55.33 | 58.22 | -42.08 |
| ICU utilization (%) | 41.81 | 47.81 | 53.82 | 34.68 | 34.95 | 35.22 | 12.86 |
| SDU utilization (%) | 53.08 | 57.73 | 62.37 | 71.41 | 71.81 | 72.22 | -14.08 |

**Table 16.** Average service performance per admitted patient.

| Performance measures | Policy 1 |  |  | Policy 2 |  |  | diff |
| --- | --- | --- | --- | --- | --- | --- | --- |
|  | Low 95% | Av | Up 95% | Low 95% | Av Total | Up 95% |  |
| Rejected Cost | 0 | 0 | 0 | 0 | 0 | 0 | 0 |
| Overstay Cost | 0.52 | 0.90 | 1.09 | 4.91 | 5.16 | 5.39 | -50.25 |
| Discharge Cost | 12.01 | 12.29 | 12.66 | 12.51 | 12.51 | 12.54 | -0.22 |
| Pre-Step-Down Cost | 0 | 0 | 0 | 37.66 | 37.73 | 37.78 | -37.73 |
| Total Cost | 13.18 | 13.57 | 13.91 | 55.07 | 55.40 | 55.70 | -41.83 |
| Admission Reward | 100 | 100 | 100 | 100 | 100 | 100 | 0 |
| step-down Reward | 24.84 | 25.11 | 25.41 | 13.18 | 13.20 | 13.24 | 11.91 |
| Total reward | 124.98 | 125.24 | 125.56 | 113.28 | 113.28 | 113.28 | 11.97 |
| Net benefit | 111.06 | 111.67 | 112.40 | 57.57 | 57.90 | 58.20 | 53.77 |

A similar analysis was performed by investigating increasing the arrival rate of patients into the ICU. The results are summarized graphically in the following figures. The blue vertical line represents, (*λ* = *μ*), the point where the arrival rate is equal to the service rate. Policy 1 is plotted in green, and Policy 2 is plotted in black.

Figure 9 shows the average number of ICU requests made in the last four months of the simulation under various arrival rates with 95% confidence interval. The confidence interval is tiny and imperceptible. As expected, ICU demand grows linearly with an increasing rate of arrival.

**Figure 9.**
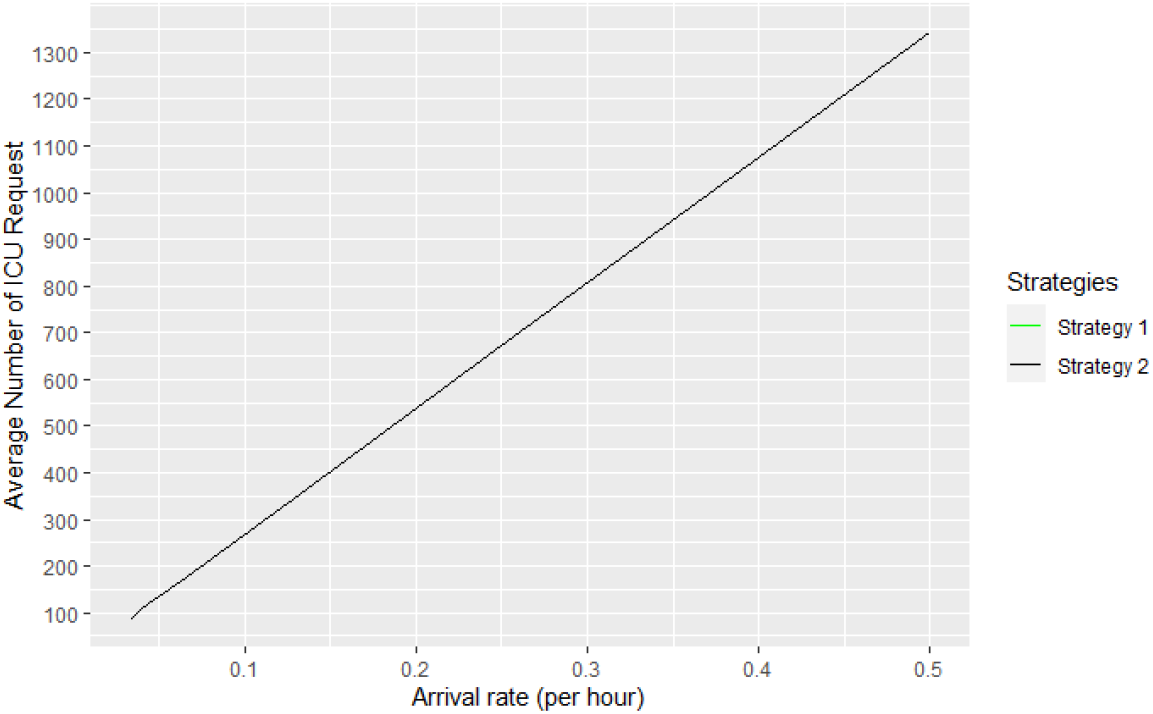
Average ICU requests

Figure 10 shows the percentage of ICU requests admitted to the ICU during the last four months of the simulation under various arrival rates with 95% confidence interval. The confidence interval is also indiscernible. When the arrival rate is less than the service rate, all patients are admitted, once the arrival rate becomes higher, we observe a linear decrease occurs in the percentage admitted into the ICU.

**Figure 10.**
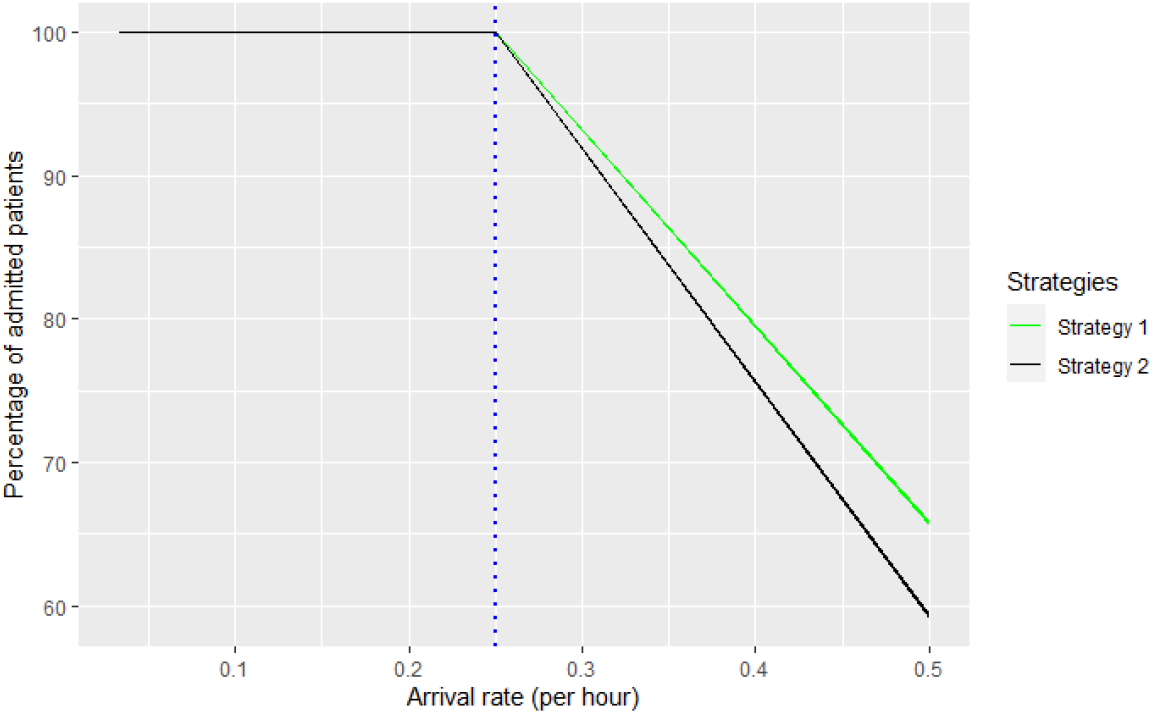
Percentage ICU Admissions versus increasing arrival rate with 95% CI. The blue vertical line represents, (*λ* = *μ*), the point where the arrival rate is equal to the service rate. Policy 1 is plotted in green, and Policy 2 is plotted in black.

Figure 11 shows the percentage of ICU requests rejected at the ICU during the last four months of the simulation under various arrival rates with 95% confidence interval. When the arrival rate is less than the service rate, no patient is rejected, once the arrival rate becomes higher, a liner increase in the percentage of rejected patients is observed.

**Figure 11.**
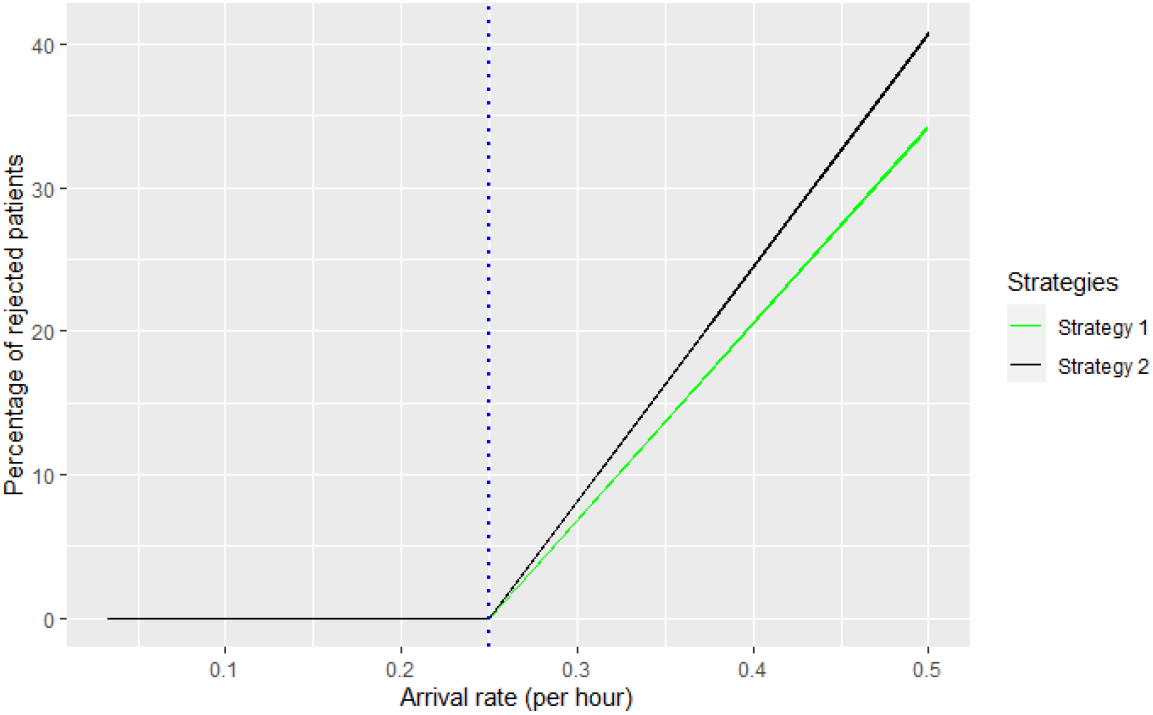
Percentage ICU rejectionsversus increasing arrival rate with 95% CI. The blue vertical line represents, (*λ* = *μ*), the point the arrival rate is equal to the service rate follows. Policy 1 is plotted in green and Policy 2 in black.

Figure 12 shows the percentage of admitted ICU patients who were stepped down under Policy 2 during the last four months of the simulation under various arrival rates with 95% confidence interval. Figure 13 shows the the percentage of admitted ICU patients who were prematurely stepped down under Policy 2 during the last four months of the simulation under various arrival rates with 95% confidence interval. When the arrival rate is less than the service rate, a little more than half the admitted patients are normally stepped down, and the rest prematurely stepped down. When the arrival rate is higher than the service rate, premature step-down leaves room for normal step-downs.

**Figure 12.**
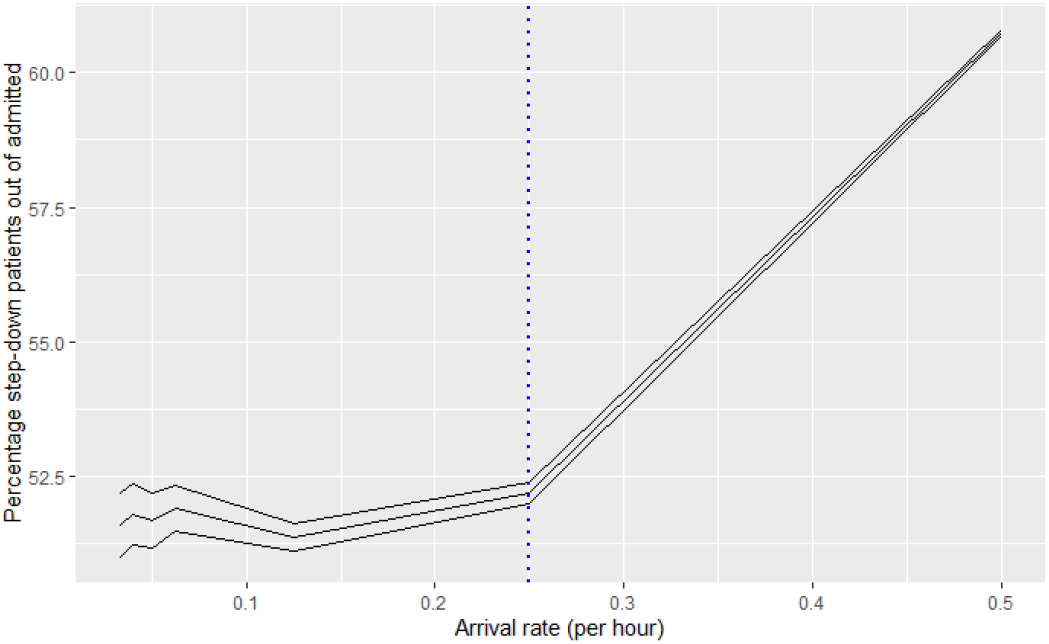
Percentage ICU step-downs using Policy 2 with 95 % CI. The blue vertical line represents, (*λ* = *μ*), the point the arrival rate is equal to the service rate.

**Figure 13.**
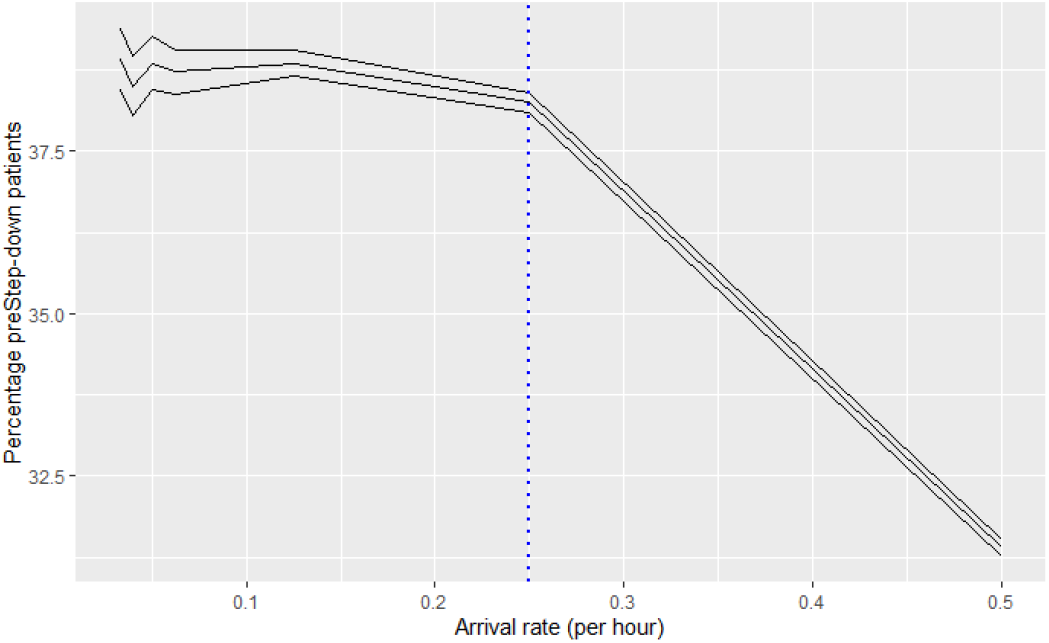
Percentage of ICU premature step-downs using Policy 2 with 95% CI. The blue vertical line represents, (*λ* = *μ*), the point the arrival rate is equal to the service rate.

Figures 14 and 15) are the average ICU and SDU utilization with 95% confidence interval. In the ICU, Policy 2 has a linearly increasing utility that is unaffected by the steady-state condition. Policy 1’s utility however, increases at a reduced upward slope when arrival rates are greater than service rate. In general, Policy 1’s ICU utility is higher than that of Policy 2. In the SDU, both policies have reduced utility steepness when arrival rates are greater than service rate. In general, Policy 2’s SDU utility is higher than that of Policy 1. ICU utility and SDU utility in Policy 1 have equivalent trends whereas SDU utility in Policy 2 is exceptional.

**Figure 14.**
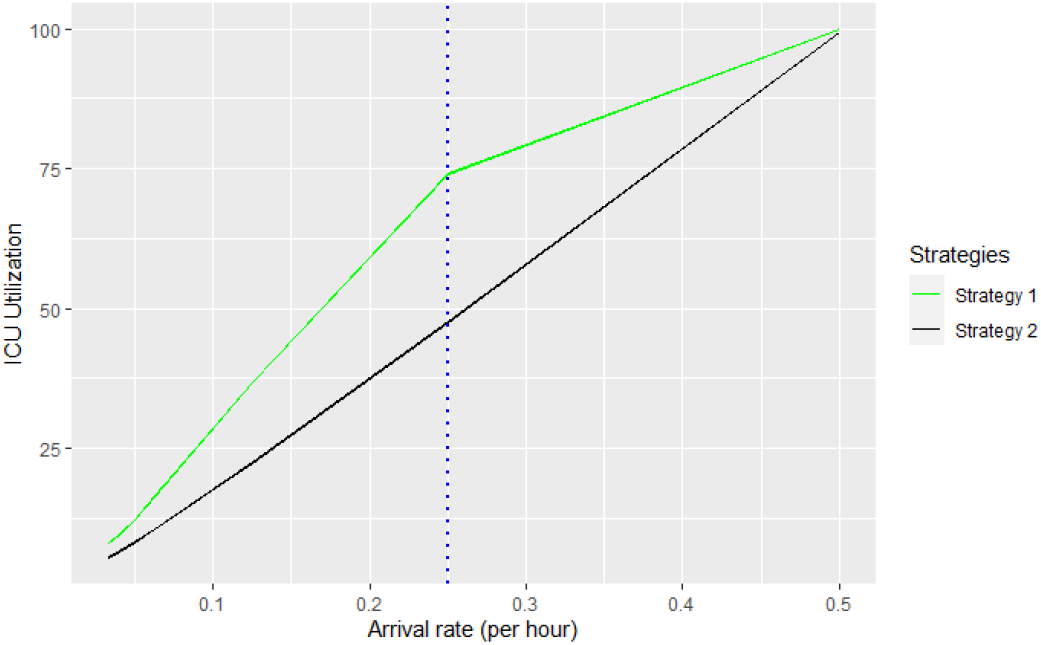
ICU utility versus increasing arrival rate with 95% CI. The blue vertical line represents, (*λ* = *μ*), the point where the arrival rate is equal to the service rate. Policy 1 is plotted in green and Policy 2 in black.

**Figure 15.**
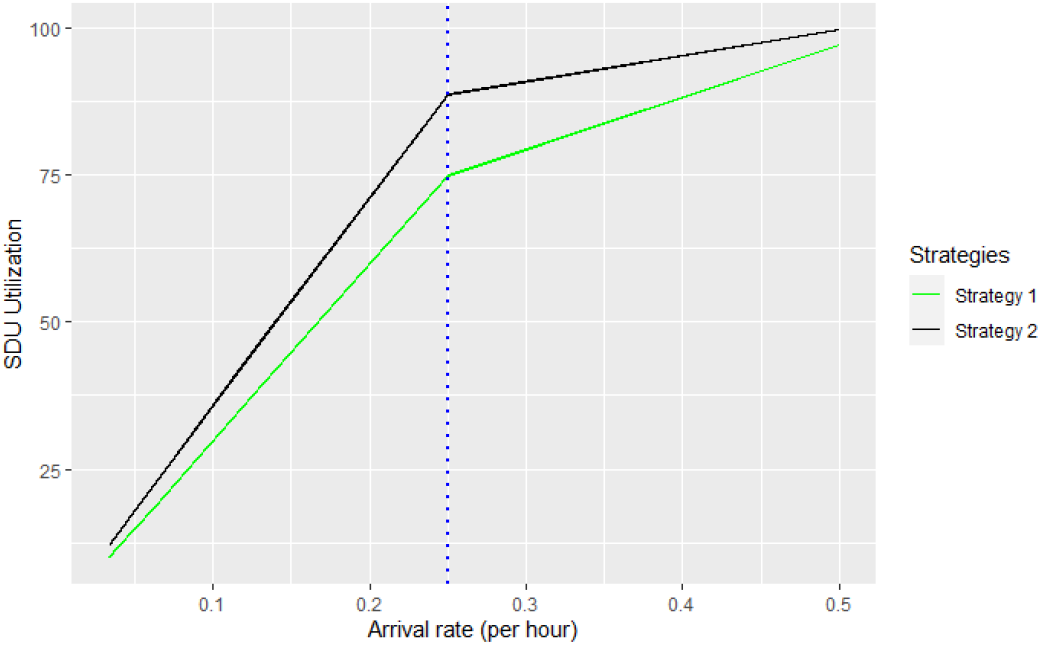
SDU utility versus increasing arrival rate with 95% CI. The blue vertical line represents, (*λ* = *μ*), the point where the arrival rate is equal to the service rate follows. Policy 1 is plotted in green while Policy 2 in black.

Figure 16 shows the average benefit per patient admitted. Both policies experience a decreasing trend with a higher steepness when the arrival rate is high. In general, Policy 1 has a higher benefit per admitted patient than Policy 2.

**Figure 16.**
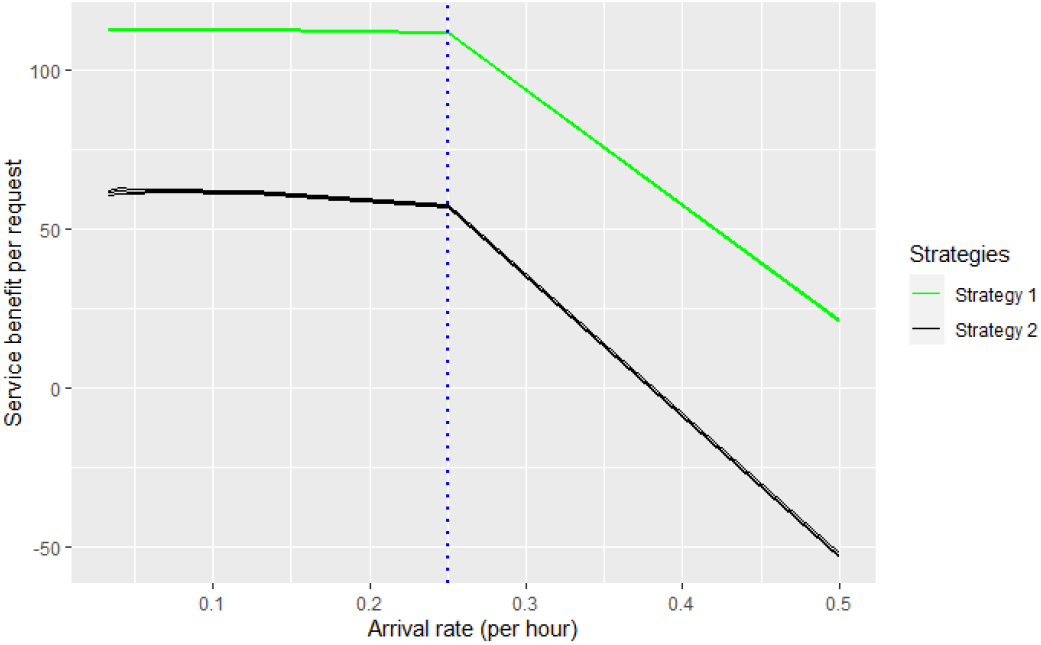
Average benefit per patient admitted with 95% CI.

## 6. Discussion

In the literature, use of an SDU has been shown to considerably increase ICU throughput (Mathews and Long 2015; Lekwijit et al. 2020; Mathews et al. 2012; Rodrigues et al. 2018). However, the effect of premature step-down has not been compared to that of rejection in a high-demand system. Two decision rules have been used to investigate the last bed problem of patient flow management in the congested environment. The sequential optimal solution stipulates admission whenever possible, i.e., if there is a mean of stepping down a patient to the SDU, either by normal or premature stepping down, it should be done, reject whenever the ICU is full, the demand is high, and the cost of premature discharge from the ICU is increased. Whenever both the ICU and the SDU are full, there is first a discharge action before a step-down action. If the cost of premature discharge is increased, then discharging patients becomes so costly that it will be rarely done, a congested ICU that prevents admission and makes other actions impossible. The interdependence of the ICU and the SDU is clearly shown in this relationship. Because the costs were assumed to be lesser than the reward, the results are robust. As expected once the cost of premature step-down is greater than the reward of admission, the optimal action is to “do nothing”, that is, it is preferable to reject arriving patients than prematurely discharge a current high-acuity patient.

Because ICU lengths of stay may be long and Policy 2 has the default action as premature step-down, it initially tends to perform excessive premature step-downs. This not only is detrimental to the health service benefit of those patients but also seems to increase the SDU length of stay in the long run. Therefore, in the long run, the SDU becomes full, and further normal step-downs are impossible. The flow rate into the SDU/ICU system becomes highly reduced. Figure 10 and Table 15 show that when the rate of arrival is less than the service rate, both policies admit on average a comparable number of patients, with Policy 1 having a higher variation due to rejections. Premature step-down causes congestion downstream. Increasing the number of patients that the SDU receives lengthens patients’ stays at the ICU, creating a worsening congestion trend, and preventing upstream patients from being moved out. When the SDU is congested, this creates overstay at the ICU, even though the ICU may seem less busy, with empty beds. Empty beds are costly to the ICU because the ICU bed capacity is hard to change whether or not the beds are used (Halpern and Pastores 2010).

Premature step-downs artificially increase the number of patients in the SDU, creating longer overstays at both the ICU and the SDU and therefore reducing the number of normal step-downs. This creates congestion at the ICU and in the whole system creating more rejection (Figure 11). Using Policy 2, increases the number of premature step-downs as the arrival rate increases, and SDU utilization increases faster than ICU utilization. An increase in SDU occupancy increases SDU overstays, prevents ICU normal and premature step-downs and causes increased ICU rejection. This partially explains the fewer admissions by Policy 2 compared to Policy 1 when arrival rates increase. An increase in rejections leads to a drop in net survival. With an arrival rate of more than half a patient per hour, whatever the policy or the system, there is no further improvement in the admittance of newly arriving patients because the ICU is already full. In such states, the system’s capacity is near 100% utilization. In general, even with premature discharge, the system will reach a point under heavy traffic where the number of rejected patients, the ICU utilization and the SDU utilization under both policies will converge as the rate of arrival increases. In these busy states, although the policy without premature step-down rejects more patients, the policy with premature step-down rejects nearly as many and performs even more premature step-downs.

Even if a patient may be denied access to the ICU because it is full, nonetheless hospital management may be pressed to identify an individual for premature step-down to prevent rejection. The perceived high risk of rejecting ICU patients and ethical considerations when rejecting a patient, constrain the practice of premature step-downs. Instead of thinking about rejection as onsite rejection of patients, rerouting of ambulance can prevent the negative impact on those patients who may be rejected and for whom a lack of bed may prove fatal. Even if Policy 2 seems right and ethical overall, under high demand, it proves more detrimental than Policy 1. Likewise, other arrangements must be considered when alternate levels of care are not available to patients and all beds are occupied.

## 7. Conclusions

This paper is concerned with the modelling of an ICU supplemented by a Step-down Unit (SDU) to assist with efficient patient flow as patients recover, in a congested environment. The Nine Equivalents of Nursing Manpower Score (NEMS) dataset for the ICU served here as the measure of patient recovery over time. The optimal actions in the resulting MDP were approximated to enable patient flow under two cases. Both cases allowed the admission of new high-acuity patients, the stepping down of existing high-acuity patients to the SDU, and the discharge of low-acuity patients from the ICU. The latter case also allowed for premature stepping down into the SDU of high-acuity patients who had not yet completed the care they would ordinarily receive in the ICU.

Numerical comparison revealed that the optimal policy for the latter case ended up performing more premature step-downs to admit a few more arrivals, relative to the case where premature step downs were not allowed. In this way, premature step-downs were shown to impact future arrivals due to the higher level of occupancy downstream, which impedes the movement of recovering patients to the SDU.This study established that NEMS works well as a proxy for classifying daily patient health states and as such, translates well into a transition matrix to be used in an MDP model. As defined in this paper, cohort health service benefit seems to be a good measure of the overall pressure on a hospital in its acute units(ICU and SDU). The rewards and costs accumulated into the health service benefit were also found to be relatively insensitive; this means that, considerable changes in the values of the rewards and costs are needed to change the general findings. In most cases, it was observed that premature step-downs stress the acute care pathway and lead to further congestion downstream. In steady-state systems with lower utilization rates, Policy 1 is recommended, with the use of an alternative level of care when there is an empty bed in the SDU in case the ICU becomes full. Surprisingly and counter-intuitively, in prolonged busy states (high utilization with high-demand scenarios), the findings of this study recommend Policy 1. This policy does not allow premature step-downs and achieve similar levels of overall health service benefit performance. The added benefit of Policy 1 is that it does so without additional stress downstream that would further impact future arrivals.

The proposed model has its limitations. This study has looked at the system level, not the individual level. Due to its intensive level of care, the individual benefits from overstaying at the ICU, but the system under-performs. If an individual overstays in the ICU, his health service benefit does not decrease, but, the system suffers a dis-utility. Especially, if a new arrival finds the ICU full. Second, NEMS is mostly used in Canada, so other jurisdictions may need to rely on other daily metrics to determine a patient’s acuity such as APACHE (all versions) and SOFA. Furthermore, the MDP model captures only the congestion zone of the system’s capacity. In a model with full capacity, the state space increases rapidly, making it less tractable and harder to solve both computationally and analytically. Moreover, the ICU/SDU ratio used in the proposed MDP model is fixed at 2:1. This was done to help formulate and solve the MDP. Finally, the health service benefit as defined in the paper may be an over-simplification of real-life phenomena in the ICU.

For future work, an analysis focused on the individual level, as opposed to the overall health service benefit of the system should be considered. Increasing the patient’s acuity level (e.g. low, medium, high NEMS), so long as this proves to be tractable, will be of great interest. Furthermore, when addressing fixed capacity ratios, one may also formulate the acute care patient flow as a queuing/capacity optimization problem. Finally, the measures of utility or dis-utility for alternative services and other kinds of ICU performances should be explored, especially as concerns the problem of the last ICU bed.

## Data Availability

The data for this study is available from the Critical Care Information System

## Acknowledgements

This research was supported by an NSERC grant. We would like to acknowledge the anonymous reviewers for commenting on earlier versions of this paper.

## Notes

### Competing Interest Statement

The authors have declared no competing interest.

### Author Declarations

Research Ethics Review Committee at King's University College.

